# Interpretable photoacoustic phenotyping of distal microcirculation for peripheral artery disease diagnosis with exploratory perioperative assessment

**DOI:** 10.64898/2026.09.02.26361270

**Authors:** Handi Deng, Tianhao Yuwen, Zipeng Li, Jiaxuan Xiang, Yizhou Bai, Naiyue Zhang, Wubing Fu, Xiaojun Wang, Jianming Guo, Weiwei Wu, Cheng Ma, Ming-Yuan Liu

## Abstract

Peripheral artery disease (PAD) spans a continuum from large-vessel obstruction to distal microvascular dysfunction, yet routine non-invasive tests, including the ankle-brachial index (ABI), do not provide structurally resolved assessment of the foot microvascular bed and may be unreliable in the setting of medial arterial calcification or perioperative follow-up. Here we developed a clinic-oriented multispectral compound-scanning photoacoustic tomography system (MCPATS) for compression-free distal toe imaging, and an interpretable photoacoustic tomography distal microcirculation score, termed PACT-DMS, for phenotyping PAD-related distal vascular abnormalities. PACT-DMS was derived from anatomically standardized distal toe sections and integrated seven prespecified vascular features spanning trunk-vessel morphology, microvascular distribution and pulsation-related dynamics through a traceable linear support vector machine. In a prospective single-centre cohort of 45 participants, the bilateral fusion PACT-DMS model distinguished patients with PAD from healthy controls with an area under the receiver operating characteristic curve of 0.964 (95% CI, 0.907–1.000) and an accuracy of 91.1% (95% CI, 82.2%–97.8%) under subject-level leave-one-out cross-validation, supported by complementary robustness analyses. Exploratory analyses further showed that PACT-DMS identified abnormal distal vascular phenotypes in 6 of 9 clinically diagnosed PAD limbs with non-abnormal ABI and visualized distal vascular-bed changes before and after revascularization. These findings support MCPATS-enabled interpretable photoacoustic vascular phenotyping as a candidate adjunctive approach for distal microcirculatory assessment in PAD; larger multicentre studies with external validation and prespecified analysis protocols will be required to define its clinical role.

## Introduction

Peripheral artery disease (PAD) is a chronic atherosclerotic disorder that impairs lower-limb perfusion in more than 200 million people worldwide and represents one of the leading vascular disease burdens after coronary and cerebrovascular disease[1–4]. PAD reflects diffuse vascular injury beyond the affected limb and is associated with increased risks of myocardial infarction, ischaemic stroke and vascular death. In patients with diabetes, medial arterial calcification or chronic ischaemia, PAD may remain clinically silent or minimally symptomatic until tissue loss or ulceration occurs; once foot ulceration coexists with PAD, long-term outcomes are markedly adverse[5–7]. Objective, reproducible and structurally interpretable assessment of the distal microvascular bed, both before clinical decompensation and during peri-procedural follow-up, therefore remains an unmet need in PAD management. Current non-invasive assessment of PAD combines macrovessel anatomical imaging with pressure– and oxygenation-based functional tests. Ultrasound, computed tomography angiography and magnetic resonance angiography can delineate stenosis, occlusion and flow abnormalities, but they do not resolve the terminal microvasculature of the foot and may be constrained by operator dependence, contrast-related risks, scan duration or motion artefacts[8–13]. The ankle-brachial index (ABI) is widely used as a first-line screening test, but can be falsely normal in patients with diabetes, medial arterial calcification or distal microvascular-predominant disease[14–17]. Transcutaneous oxygen pressure (TcPO₂) can estimate wound-healing potential, but is time-consuming and sensitive to local skin and ambient conditions[18–22]. These tests therefore primarily index large-vessel patency, ankle-level haemodynamics or indirect tissue oxygenation, but do not provide standardized, spatially resolved and structurally interpretable phenotyping of the distal foot microvasculature.

The distal microvascular bed is increasingly recognized as a clinically relevant determinant of PAD progression and treatment response. PAD pathophysiology extends beyond large-artery stenosis to include endothelial dysfunction, arteriolar remodelling, capillary rarefaction, altered branching topology and spatial heterogeneity in haemoglobin distribution and oxygenation[23–31]. These abnormalities may precede typical symptoms or measurable declines in resting ABI[32–35]. Even after revascularization restores upstream arterial patency, wound healing, pain relief and long-term limb outcomes depend on whether tissue-level reperfusion and microcirculatory recovery are achieved[36–44]. Direct characterization of distal vascular-bed structure, distribution and dynamic behaviour may therefore provide a phenotype dimension complementary to conventional pressure-based indices, with potential value for PAD stratification and perioperative assessment.

Photoacoustic imaging (PAI) combines haemoglobin-based optical absorption contrast with ultrasound-mediated detection depth, enabling contrast-agent-free, spatially resolved vascular imaging[45–54]. In lower-limb and foot vascular disease, multispectral optoacoustic imaging has been used to assess calf-muscle oxygenation gradients associated with PAD stage, walking performance and imaging-based classification[55, 56], whereas three-dimensional foot photoacoustic tomography has improved visualization of dorsal and plantar vascular structures by expanding field-of-view coverage and reducing limited-view artefacts[56–59]. More recently, radiomic analysis of photoacoustic foot images has been explored for predicting perfusion and vascular-bed function[60]. Beyond PAD-specific studies, raster-scan optoacoustic mesoscopy (RSOM) has enabled label-free quantification of diabetes-related skin microangiopathy through morphophysiological cutaneous features and optoacoustic biomarkers of dermal microvascular density and skin microanatomy[61, 62]. Collectively, these studies support PAI and RSOM as quantitative tools for characterizing superficial peripheral vascular structure, perfusion-related status and microvascular remodelling associated with PAD and metabolic microangiopathies[63,64]. However, an interpretable, physiologically grounded scoring framework for PAD-oriented distal vascular phenotyping remains lacking, and the complementary value of photoacoustic measures relative to routine clinical indices such as ABI, as well as their potential utility for perioperative assessment, remains insufficiently defined.

Here we developed a multispectral compound-scanning photoacoustic tomography system (MCPATS) for compression-free imaging of the distal foot and toe-pulp vasculature, and established PACT-DMS as an interpretable photoacoustic scoring framework that converts anatomically standardized distal toe images into physiologically labelled vascular phenotypes for PAD assessment. In this study, distal microcirculation is defined operationally as the PACT-resolved vascular bed within the distal foot region, including longitudinal trunk vessels, branch vessels and small-vessel networks that reflect terminal perfusion-bed structure and dynamics. From anatomically standardized toe sections, PACT-DMS extracts seven prespecified semantic vascular features spanning trunk-vessel morphology, microvascular distribution and pulsation-related dynamics, and integrates them through a traceable linear support vector machine. We evaluated this system-and-framework approach in a prospective single-centre cohort of patients with PAD and age-matched healthy controls, and further explored its utility in clinically diagnosed PAD limbs with non-abnormal ABI and in perioperative distal vascular-bed assessment. Rather than replacing ABI, computed tomography angiography, ultrasound or other established vascular tests, MCPATS-enabled PACT-DMS is proposed as a candidate adjunctive approach for structurally resolved distal vascular phenotyping, with potential clinical value in PAD assessment, non-abnormal ABI presentations and perioperative evaluation of distal vascular-bed status.

## Results

### MCPATS-enabled standardized distal foot imaging in a prospective PAD cohort

To support PAD-oriented distal vascular phenotyping in human participants, we developed a photoacoustic tomography system tailored for compression-free imaging of the distal foot vasculature, termed MCPATS. The system was designed to meet three practical requirements: stable visualization of small superficial vessels, repeatable imaging of anatomically corresponding toe regions and flexible acquisition of three-dimensional structural, multispectral and dynamic vascular information under standardized coupling conditions. Functionally, the compound-scanning design supports multi-angle acquisition for more isotropic three-dimensional vascular imaging, as well as rapid single-angle three-dimensional scanning when acquisition efficiency is prioritized; foot-shape-informed scanning-path planning further supports individualized coverage of the distal foot and toes. Together with temperature-controlled water coupling, foot stabilization and energy-normalized photoacoustic acquisition, this configuration was intended to reduce pressure-induced vascular distortion and support repeatable assessment of distal vascular morphology and pulsation-related dynamics. Detailed system architecture, imaging parameters and acquisition procedures are described in Methods.

We then deployed MCPATS in a prospective single-centre observational study that included 24 clinically confirmed patients with PAD and 21 age-matched healthy volunteers, all of whom underwent three-dimensional toe PACT imaging. The imaging protocol translated the system design into a standardized clinical workflow: participants were imaged in a seated position, with warm-water coupling at 30–35 °C, foot stabilization and avoidance of lower-limb compression by clothing or posture, to minimize the influence of posture, temperature and external pressure on peripheral vascular status. Based on integrated assessment of clinical diagnosis, symptomatic side and vascular imaging data, including CTA or digital subtraction angiography, both lower limbs of the enrolled patients with PAD showed varying degrees of PAD-related vascular involvement. Therefore, in limb-level analyses, both feet of patients with PAD were included as PAD observations, whereas both feet of healthy volunteers were included as healthy-control observations. After exclusion of limbs with insufficient signal quality, reconstruction quality or invalid limb status, 44 PAD limbs and 40 healthy limbs were included in the limb-level analysis. Figure 1 summarizes the overall MCPATS-enabled study workflow (Fig. 1a), PACT-DMS model construction from representative toe sections (Fig. 1b) and exploratory postoperative phenotyping scheme (Fig. 1c). Demographic and clinical characteristics of the participants are summarized in Supplementary Table 1, and de-identified individual-level baseline data are provided in Supplementary Data 1.

**Fig. 1.**
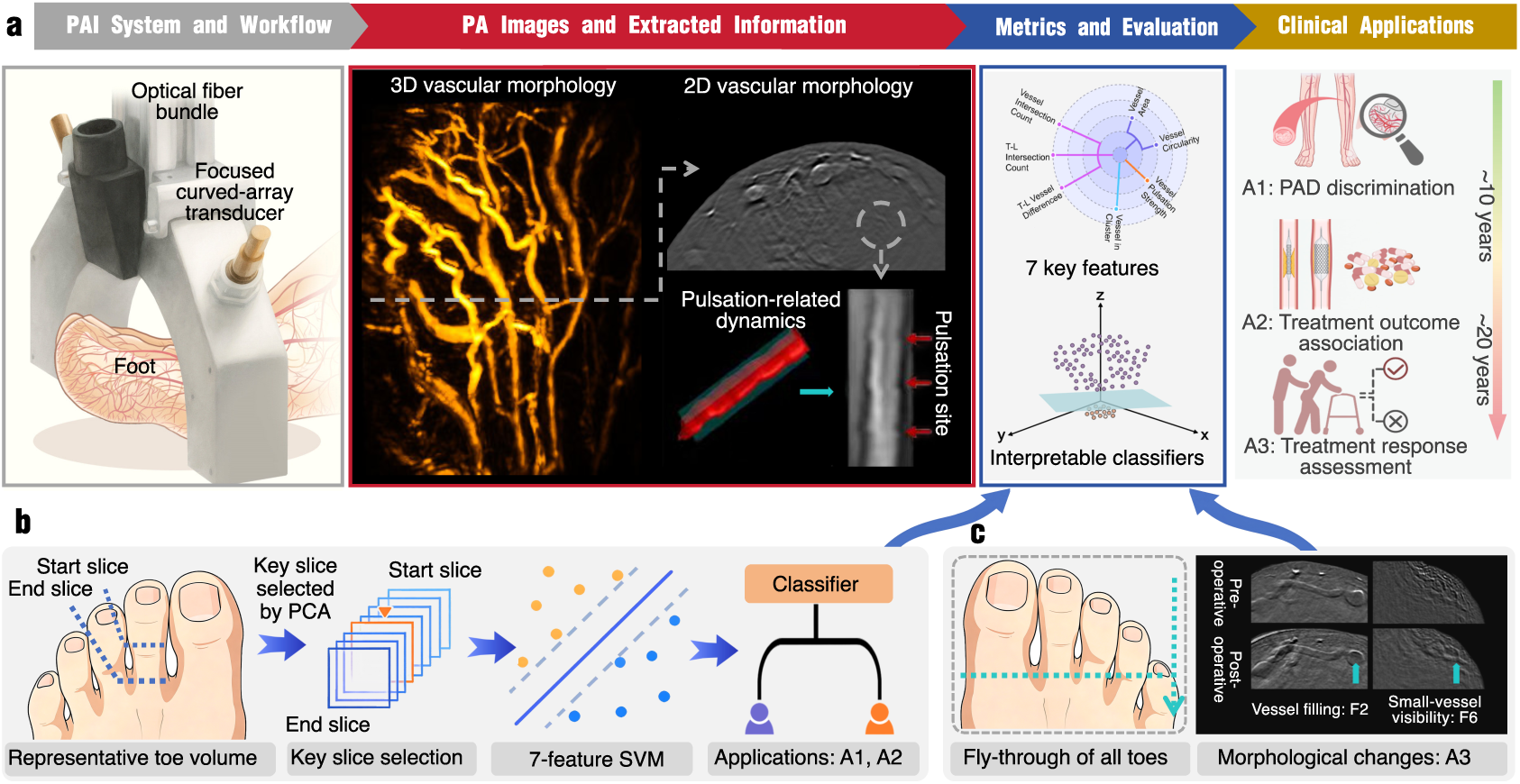
| MCPATS-enabled PACT-DMS framework for distal vascular phenotyping and clinical evaluation in PAD. **a**, System and analytical workflow. The multispectral compound-scanning photoacoustic tomography system (MCPATS) enabled standardized three-dimensional imaging of the distal toe vasculature, extraction of vascular morphology and pulsation-related dynamics, construction of the photoacoustic tomography distal microcirculation score (PACT-DMS), and evaluation across three clinical applications. **b**, PACT-DMS development. A representative second-toe volume was defined between the start and end slices, and principal component analysis (PCA) was used to select a key slice from serial PACT sections. Seven interpretable vascular features were integrated using a linear support vector machine (SVM) to generate traceable PACT-DMS decision scores. The PACT-DMS framework was evaluated for PAD discrimination (A1) and exploratory treatment-outcome association (A2). **c**, Workflow for exploratory treatment-response assessment. Fly-through visualization across all toes, together with paired preoperative and postoperative PACT images, was used to characterize distal vascular morphological changes after lower-limb revascularization, including changes in vessel filling and small-vessel visibility. These postoperative observations were linked to the corresponding PACT-DMS feature dimensions F2 and F6 and used for treatment-response assessment (A3). A1–A3 denote three applications: PAD discrimination, treatment-outcome association and treatment-response assessment, respectively. F2 and F6 denote L-vessel circularity and S-vessel count, respectively.

### PACT-DMS converts standardized toe images into interpretable distal vascular phenotypes

We next developed PACT-DMS as an interpretable distal vascular phenotyping and scoring framework that transforms standardized toe PACT images into physiologically labelled vascular features and traceable decision scores. The first step in this transformation was to define a standardized image input for cross-participant feature extraction. Therefore, the second toe was selected as the primary analysis region, and a representative section was selected within a predefined anatomical segment using a PCA-assisted procedure. This strategy was designed to reduce the dimensional burden caused by three-dimensional anatomical variation and redundant slices while preserving the dominant vascular structural pattern; it was not intended to imply that a single two-dimensional section is intrinsically superior to whole-volume analysis. The anatomical rationale for selecting the second toe and the representative-section selection workflow are shown in Supplementary Fig. 7, and adjacent-section consistency is analysed in Supplementary Fig. 8.

Representative PACT cross-sections showed visually distinct distal vascular patterns between healthy participants and patients with PAD (Fig. 2a). Healthy participants typically showed continuous trunk vessels, better vessel filling and relatively uniform small-vessel distribution, whereas patients with PAD showed varying degrees of reduced distal trunk-vessel visibility, irregular vascular morphology, disordered small-vessel distribution or local microvascular remodelling. Based on joint interpretation by vascular surgeons and photoacoustic imaging engineers, vascular structures in the representative sections were assigned to three clinically interpretable morphological categories (Fig. 2b): L-type vessels, defined as longitudinal trunk vessels running along the long axis of the toe and representing the main distal perfusion conduits; T-type vessels, defined as transverse or oblique branches extending from longitudinal trunks and reflecting branch or collateral-like connecting structures; and S-type vessels, interpreted as PACT-visible small-vessel signals or small-vessel clusters associated with the distal microvascular bed, rather than direct single-capillary visualization.

**Fig. 2.**
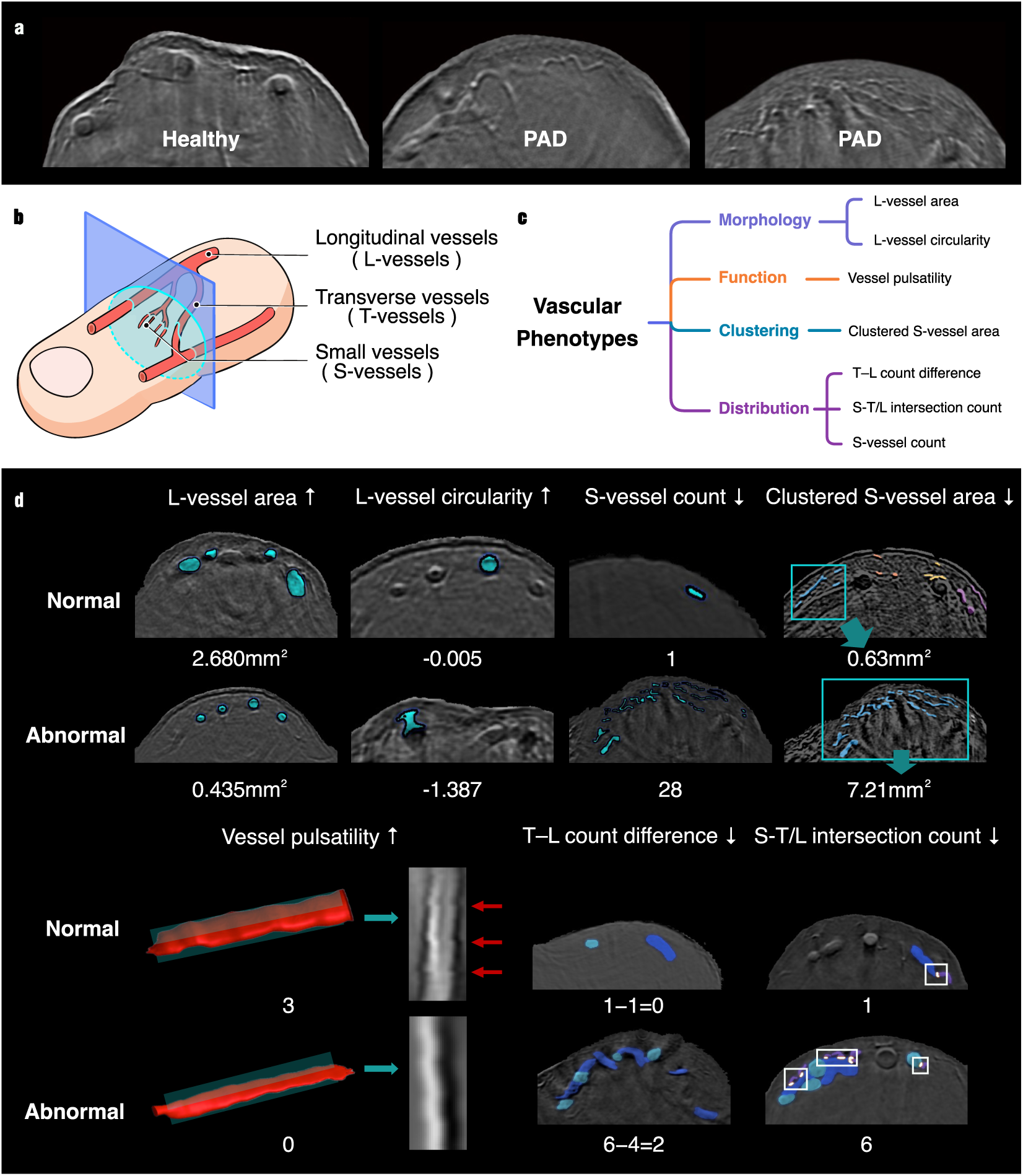
| L/T/S vascular semantic framework and seven-feature construction of PACT-DMS. **a**, Representative PACT cross-sections from healthy participants and patients with PAD, showing PAD-related distal vascular alterations, including reduced trunk-vessel visibility, luminal narrowing and increased morphological irregularity. **b**, Schematic definition of three vascular structure categories in representative toe sections: longitudinal trunk vessels running along the toe axis (L-type), transverse or oblique branch vessels extending from trunk vessels (T-type), and discrete small-vessel signals or microvascular clusters (S-type). The blue plane indicates the anatomically standardized representative cross-section used for feature extraction. **c**, Overview of the seven semantic vascular features used to construct PACT-DMS, grouped into trunk-vessel morphology, pulsation-related dynamic behaviour, microvascular clustering and spatial distribution. **d**, Representative normal and abnormal examples of the seven features, including L-vessel area, L-vessel circularity, vessel pulsation strength, Clustered S-vessel area, T–L vessel-count difference, S-vessel count and S-T/L vessel intersection count. Pseudocolour overlays indicate vascular segmentation or detection results, and numerical values indicate representative feature measurements. Arrows indicate the direction of change in healthy controls relative to PAD patients.

On this L/T/S vascular semantic framework, we constructed seven physiologically motivated vascular features to form PACT-DMS (Fig. 2c,d). L-vessel area (F1) and L-vessel circularity (F2) describe the cross-sectional size and morphological integrity of longitudinal trunk vessels. Vessel pulsation strength (F3) describes periodic vascular displacement, morphological change or filling change in serial PACT images and was treated as an indirect perfusion-related dynamic phenotype rather than a direct measurement of flow velocity or flow volume. Clustered S-vessel area (F4) and S-vessel count (F6) characterize small-vessel aggregation and distributional burden, whereas T–L vessel-count difference (F5) and S-T/L intersection count (F7) describe vessel-type redistribution and topological coupling among trunk vessels, branches and the microvascular network. These seven features span trunk-vessel morphology, pulsation-related dynamics, microvascular clustering and spatial distribution, and were integrated using a linear support vector machine to generate a traceable PACT-DMS decision score. Feature definitions, potential pathophysiological interpretations and representative examples are shown in Fig. 2, Supplementary Figs. 5 and 6 and Supplementary Table 5; individual limb-level F1–F7 feature values are provided in Supplementary Data 3.

### Prospective evaluation of PACT-DMS for interpretable PAD discrimination

To assess the clinical relevance of the PACT-DMS framework, we evaluated whether the derived distal vascular phenotype could discriminate patients with PAD from healthy controls in the prospective cohort. Two complementary analytical settings were considered: a unilateral limb-level model, which characterized side-specific distal vascular phenotypes, and a bilateral fusion model, which integrated left– and right-foot decision scores at the participant level. To avoid optimistic bias from bilateral measurements of the same participant, model performance was evaluated using subject-level leave-one-out cross-validation, in which all available limbs from the held-out participant were excluded from model training.

The unilateral model achieved strong diagnostic discrimination, with an AUC of 0.904 and an accuracy of 83.3% (Fig. 3a–c). After bilateral fusion, performance further improved, reaching an AUC of 0.964 (95% CI, 0.907–1.000) and an accuracy of 91.1% (95% CI, 82.2%–97.8%). With PAD treated as the clinical positive class, the unilateral model achieved a sensitivity of 84.1% and a specificity of 82.5%, whereas the bilateral fusion model achieved a sensitivity of 91.7% and a specificity of 90.5%. The use of a linear support vector machine preserved the link between model output and the underlying vascular features, allowing PACT-DMS decision scores to be traced back to morphological, distributional and dynamic vascular phenotypes. Internal robustness analyses, including bootstrap uncertainty estimation, participant-level permutation testing, leave-one-feature-out ablation and covariate sensitivity analysis, are described in Methods and summarized in Supplementary Figs. 12 and 13.

**Fig. 3.**
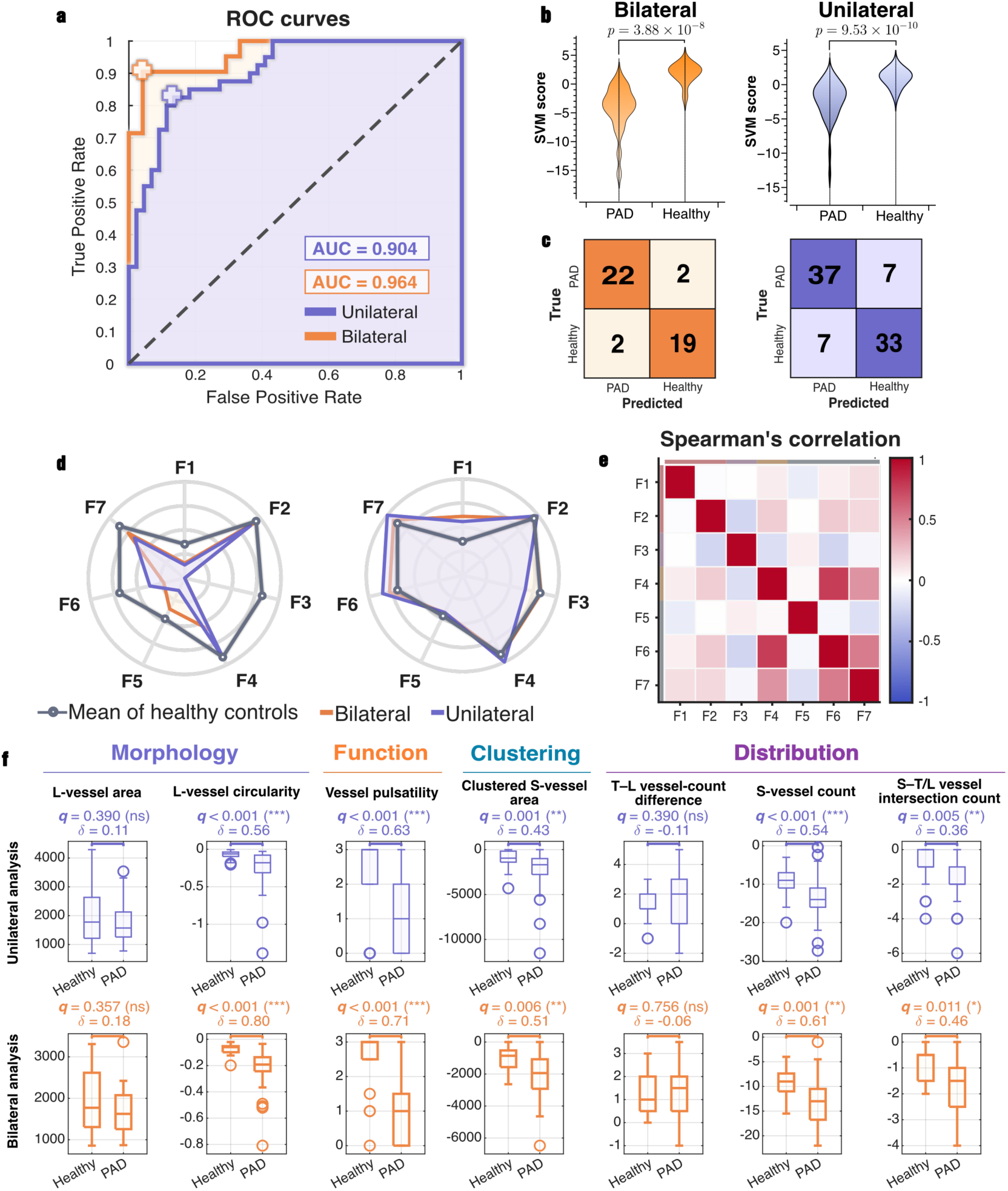
| Prospective evaluation and feature-level characterization of PACT-DMS for PAD discrimination. **a**, Receiver operating characteristic curves for the unilateral limb-level model and bilateral fusion model evaluated using subject-level leave-one-out cross-validation, in which all available limbs from the held-out participant were excluded from model training. **b**, Violin plots of linear support vector machine decision scores for the bilateral fusion and unilateral models, showing the distributions of scores in patients with PAD and healthy controls. **c**, Corresponding confusion matrices for the bilateral fusion and unilateral models, with PAD treated as the clinical positive class. **d**, Radar plots of the seven PACT-DMS features under unilateral and bilateral analyses. Feature values were normalized before radar-plot visualization and are shown relative to the mean values of the healthy controls. Representative patient profiles are shown on the left, and representative healthy-participant profiles are shown on the right. **e**, Spearman correlation matrix of the seven features in the unilateral analysis, showing relationships among the interpretable vascular feature dimensions. **f**, Box-plot comparisons of the seven PACT-DMS features in the unilateral analysis (top, purple) and bilateral analysis (bottom, orange). The features are grouped as Morphology (F1, L-vessel area; F2, L-vessel circularity), Function (F3, Vessel pulsatility), Clustering (F4, Clustered S-vessel area) and Distribution (F5, T–L vessel-count difference; F6, S-vessel count; F7, S–T/L vessel intersection count). False-discovery-rate-corrected q values and Cliff’s δ effect sizes are shown. Limb-level model outputs, F1–F7 feature values and participant-level bilateral fusion results are provided in Supplementary Data 3 and Supplementary Data 4.

Feature-level analyses further supported the interpretability of PACT-DMS. Radar plots showed coordinated deviations of PAD profiles from the healthy reference pattern, rather than isolated changes in a single metric (Fig. 3d). Spearman correlation analysis indicated that the seven features were not uniformly redundant: except for the expected correlation between Clustered S-vessel area and S-vessel count, which both describe microvascular remodelling, most feature pairs showed weak to moderate correlations (Fig. 3e). These results suggest that PACT-DMS integrates complementary information from trunk-vessel morphology, microvascular remodelling, topological coupling and perfusion-related dynamics.

Group-wise feature comparisons revealed that five of the seven features reached statistical significance after false-discovery-rate correction, including L-vessel circularity, vessel pulsation strength, Clustered S-vessel area, S-vessel count and S-T/L intersection count (F2–F4 and F6–F7; q < 0.05; Fig. 3f). In physiological terms, PAD was associated with impaired trunk-vessel morphology, weakened pulsation-related dynamics, increased small-vessel clustering and altered coupling between trunk vessels and the microvascular network. L-vessel area showed only a non-significant tendency to be lower in PAD, whereas T–L vessel-count difference captured broader interindividual heterogeneity in branch-related remodelling. Taken together, distal vascular abnormality in PAD was not simply reflected by reduced trunk-vessel size. Rather, these feature-level results support the interpretation that PACT-DMS captures a coordinated distal vascular-bed phenotype involving trunk-vessel morphology, pulsation-related dynamics and small-vessel network organization, rather than acting as a single-vessel calibre surrogate.

Together, these results indicate that PACT-DMS provides an interpretable and quantitatively discriminative representation of MCPATS-resolved distal vascular-bed abnormalities in this single-centre cohort. Although external validation is required, the combination of standardized distal foot PACT imaging, low-dimensional semantic vascular features and a linear scoring framework supports the physiological traceability of the model output and provides a transparent basis for subsequent clinical evaluation.

### PACT-DMS reveals abnormal distal vascular phenotypes in clinically diagnosed PAD limbs with non-abnormal ABI

We next examined a clinically relevant subgroup in which pressure-based assessment may be less informative: clinically diagnosed PAD limbs with non-abnormal ABI. This exploratory analysis was designed to assess whether PACT-DMS could provide complementary distal vascular-bed information in ABI-non-abnormal presentations, rather than to establish diagnostic superiority over ABI. Among 44 clinically diagnosed PAD limbs with available ABI records, 35 had abnormal ABI and 9 had non-abnormal ABI, using ABI < 0.90 to define abnormal ABI and ABI ≥ 0.90 to define non-abnormal ABI (Fig. 4a). Of these nine limbs, six were classified as PACT-positive by PACT-DMS, suggesting abnormal distal vascular phenotypes despite non-abnormal pressure-based measurements. When expressed as z-score deviations relative to the healthy-reference cohort, the feature-deviation plot and heat map showed that these abnormalities were heterogeneous rather than uniform across cases, involving different combinations of trunk-vessel morphology, small-vessel distribution, topological coupling and dynamic vascular behaviour (Fig. 4a,c). Representative PACT cross-sections further showed insufficient vessel filling, irregular trunk morphology, altered small-vessel distribution and local microvascular remodelling in these PACT-positive limbs with non-abnormal ABI (Fig. 4b). One clinically illustrative limb, Patient 2L, showed left-foot ulceration despite non-abnormal bilateral ABI, with abnormal PACT-DMS findings in the affected limb; detailed case-level information is provided in Supplementary Table 7 and Supplementary Data 2.

**Fig. 4.**
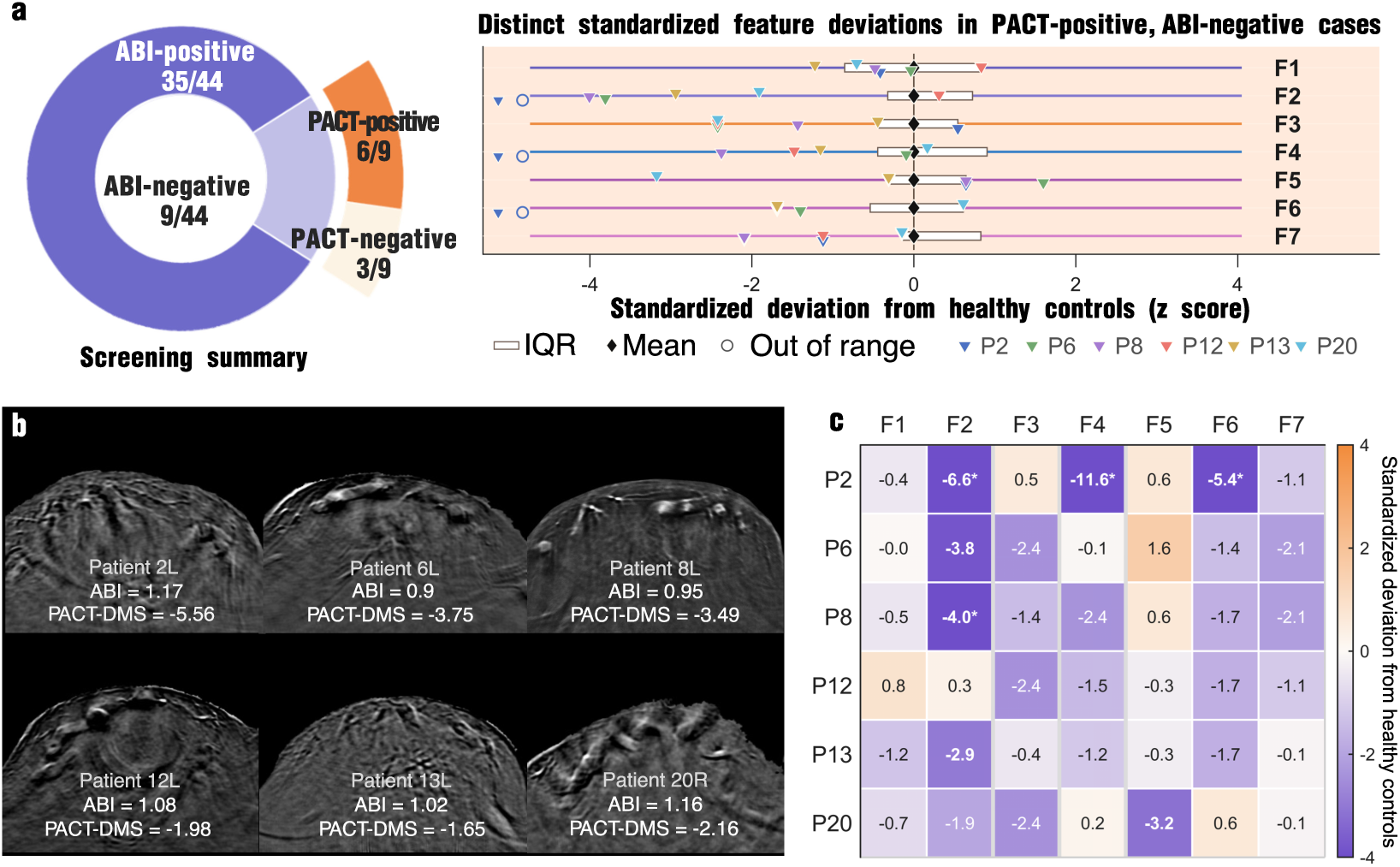
| PACT-DMS reveals heterogeneous distal vascular phenotypes in clinically diagnosed PAD limbs with non-abnormal ABI. **a**, Among nine clinically diagnosed PAD limbs with non-abnormal ABI, six were classified as PACT-DMS-positive and three as PACT-DMS-negative. The right panel shows standardized deviations (z scores) of the seven PACT-DMS features in the six PACT-positive limbs, calculated relative to the healthy-control reference cohort. Boxes indicate interquartile ranges, black diamonds indicate reference means, open circles indicate values outside the plotting range and coloured triangles indicate individual limbs. **b**, Representative PACT cross-sectional images of the six PACT-DMS-positive limbs with non-abnormal ABI, showing varying degrees of distal vascular abnormality, including insufficient vessel filling, irregular trunk-vessel morphology and altered small-vessel distribution. **c**, Heat map of standardized deviations (z scores) for the same seven interpretable vascular features in the six PACT-DMS-positive limbs. Individual limbs exhibited distinct combinations of abnormal features, indicating heterogeneous distal vascular-bed involvement despite non-abnormal ABI. Together, these exploratory findings suggest that PACT-DMS may provide complementary information on distal vascular phenotypes not fully captured by pressure-based ABI. L and R denote the left and right limbs, respectively. F1– F7 denote L-vessel area, L-vessel circularity, vessel pulsation strength, S-vessel in cluster, T – L vessel difference, S-vessel count and S – T/L intersection count, respectively. Negative and positive z scores indicate feature values below and above the healthy-control mean, respectively. Source data are provided in Supplementary Data 3, and de-identified clinical and vascular imaging summaries for the corresponding cases are provided in Supplementary Data 2.

In this exploratory nine-limb subgroup, PACT-DMS identified abnormal distal vascular phenotypes in six clinically diagnosed PAD limbs despite non-abnormal ABI. These findings provide preliminary evidence that PACT-DMS may capture MCPATS-resolved distal vascular-bed information not fully reflected by ABI and support the potential utility of photoacoustic distal vascular phenotyping as a complementary readout of vascular-bed structure and dynamic behaviour. Prospective validation in larger populations at risk of falsely normal ABI, including patients with diabetes, advanced chronic kidney disease or dialysis, is required before any clinical recommendation.

### Exploratory perioperative assessment using preoperative PACT-DMS and postoperative distal vascular phenotypes

We further explored whether PACT-DMS could extend from cross-sectional distal vascular phenotyping to perioperative assessment before and after revascularization. Although revascularization aims to restore upstream arterial patency, postoperative symptom relief, wound healing and long-term limb outcomes depend on whether distal tissue-level reperfusion and vascular-bed recovery are achieved, underscoring the importance of follow-up assessment of distal vascular status. We therefore examined whether baseline PACT-DMS reflected distal vascular-bed reserve associated with subsequent recovery, and whether PACT-DMS-derived postoperative phenotypes captured distal vascular refilling and small-vessel visibility after revascularization. Accordingly, we performed two exploratory perioperative analyses: a six-patient preoperative association analysis with 6-month Rutherford recovery, and a twelve-patient paired pre/postoperative imaging analysis focused on distal vascular refilling.

Preoperatively, six patients with PAD had complete PACT imaging and clinical follow-up after revascularization (Fig. 5). Representative operated-side or clinically corresponding toe sections and seven-feature PACT-DMS radar profiles showed interpatient heterogeneity in distal vascular involvement, including impaired trunk-vessel morphology, altered small-vessel distribution and pulsation-related abnormalities (Fig. 5a,b). For Patient 14, because the right-foot phenotype was not directly assigned a PACT-DMS score, the left-foot PACT-DMS score was used as a conservative quantitative representation, while the right-foot image was retained as qualitative evidence of severe bilateral involvement (Supplementary Fig. 14). When compared with Rutherford recovery trajectories, more PAD-like preoperative PACT-DMS phenotypes tended to accompany less favourable clinical recovery (Fig. 5c). Patient 14 showed severe distal vascular-bed depletion resembling a desert-foot-like phenotype[41] and was the only participant carrying an MTHFR 677C>T mutation, which is noted as case-level context in Supplementary Table 2 without implying causal inference.

**Fig. 5.**
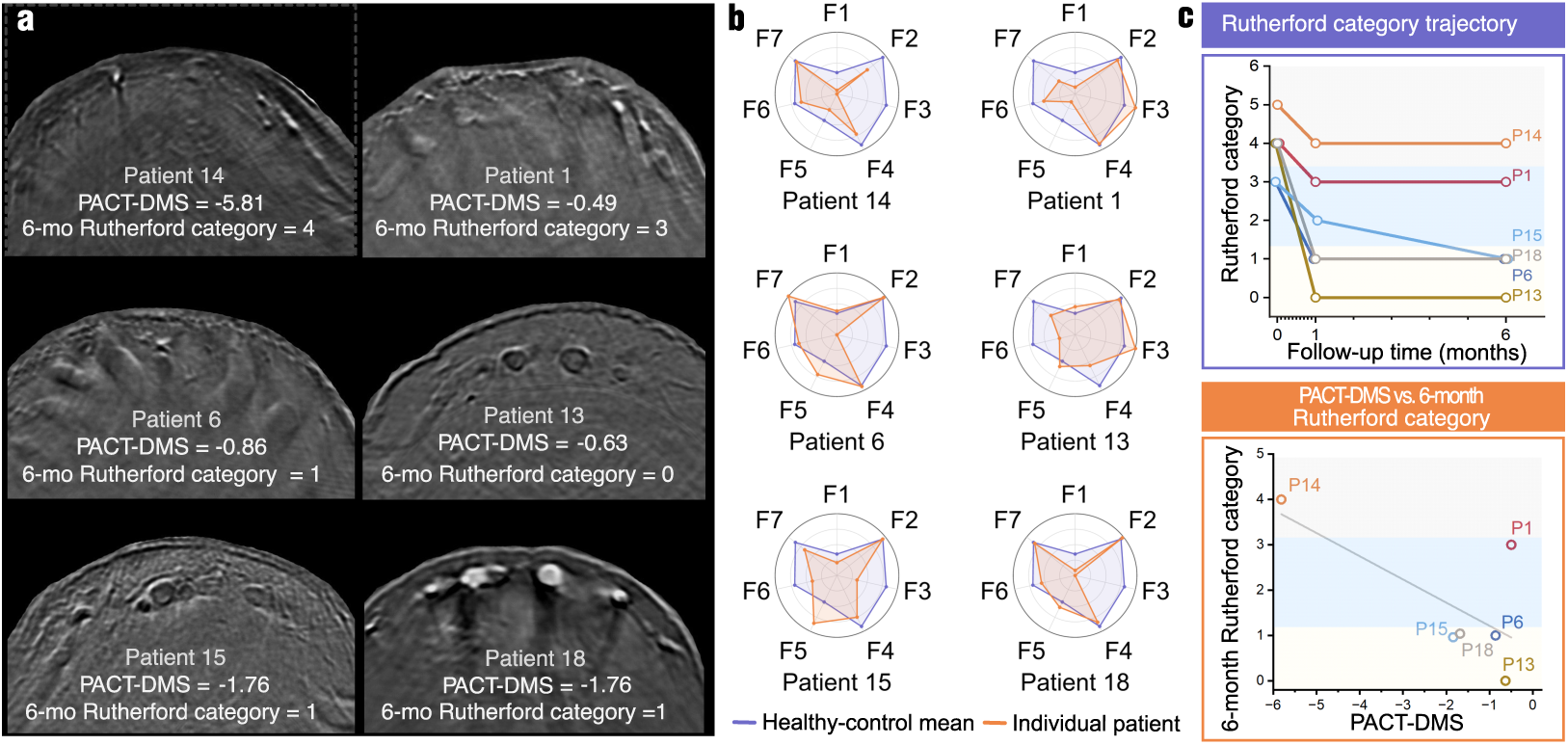
| Preoperative PACT-DMS phenotypes and exploratory recovery trends after revascularization. **a**, Representative preoperative PACT cross-sections from six patients with PAD who underwent revascularization and completed 6-month follow-up. Each image is annotated with the preoperative PACT-DMS score and 6-month Rutherford category. For Patient 14, the right-foot image is shown, whereas the left-foot PACT-DMS score was used because a right-foot score was unavailable; the rationale for this substitution is detailed in Supplementary Fig. 14. **b**, Radar plots of the seven preoperative PACT-DMS features in the same patients, referenced to the healthy-control mean, showing interpatient heterogeneity in trunk-vessel morphology, small-vessel distribution and pulsation-related features. **c**, Rutherford category trajectories and their exploratory relationship with preoperative PACT-DMS. The upper panel shows Rutherford categories before and at 1 and 6 months after revascularization; the lower panel shows the relationship between preoperative PACT-DMS and 6-month Rutherford category. Background shading provides a visual guide to favourable (categories 0–1), intermediate (2–3) and less favourable (4–6) clinical status and does not represent model-derived or statistically defined thresholds. In this small cohort, more PAD-like preoperative phenotypes tended to accompany less favourable recovery trajectories, but these observations do not establish prognostic performance. F1–F7 denote L-vessel area, L-vessel circularity, vessel pulsation strength, S-vessel in cluster, T–L vessel difference, S-vessel count and S–T/L intersection count, respectively. Source data are provided in Supplementary Data 2–4 and Supplementary Table 2.

Postoperatively, 12 patients underwent paired PACT imaging and ABI testing before and after revascularization within the same treatment cycle. Using the PACT-DMS-derived postoperative phenotyping procedure defined in Methods, we evaluated changes in distal vessel filling, vessel fullness and small-vessel visibility across serial sections and multiple toes. ΔPACT-DMS was defined as the number of imaged toes showing visible postoperative improvement relative to baseline, with higher values indicating more extensive improvement of the distal vascular bed.

Clinical follow-up indicated favourable recovery in all patients. However, ABI changes in some patients did not fully reflect the corresponding improvement in the distal vascular bed, whereas PACT-DMS-derived postoperative phenotypes consistently showed clear changes. Multi-case paired comparisons in Supplementary Figs. 15–17 demonstrated enhanced distal vessel filling, clearer visualization of previously weak vascular structures and increased small-vessel signals after revascularization. In the representative limbs shown in Fig. 6b, Patient 16R and Patient 20L showed only limited changes in ABI, with ΔABI values of 0.05 and 0.11, respectively, whereas their ΔPACT-DMS values were 2 and 5. Patient 20L underwent femoral, infrapopliteal and pedal-vessel revascularization and showed more extensive PACT phenotypic improvement than Patient 16R, who underwent femoral and infrapopliteal revascularization, qualitatively consistent with the greater distal extent of intervention.

**Fig. 6.**
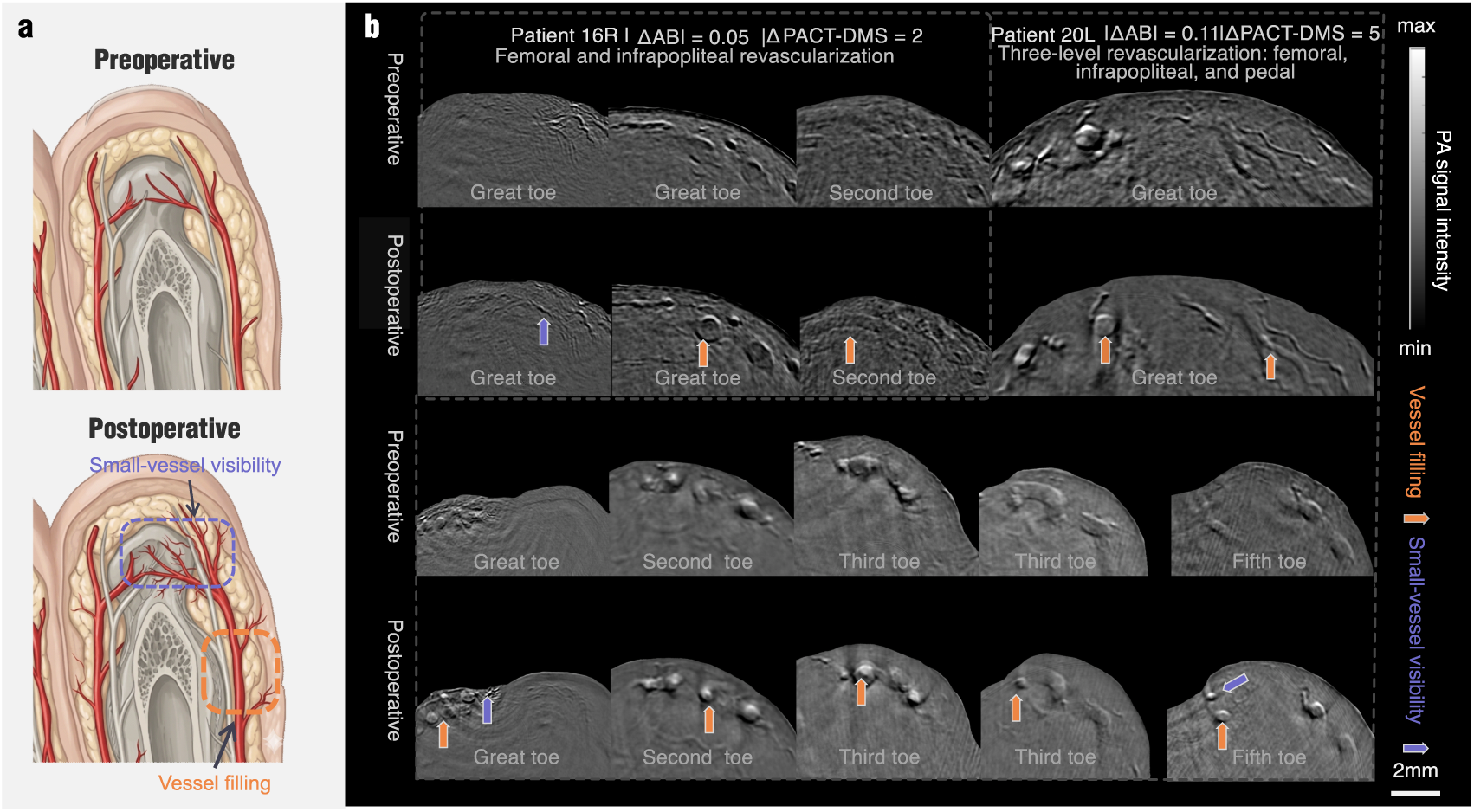
| PACT-DMS-derived postoperative phenotypes visualize distal vascular refilling after revascularization. **a**, Conceptual illustration of distal vascular changes after revascularization. Restoration of upstream arterial patency may enhance distal trunk-vessel filling, improve small-vessel visibility and re-establish local perfusion pathways. **b**, Representative paired preoperative and postoperative PACT cross-sections from two PAD limbs with limited ABI changes. Patient 16R underwent femoral and infrapopliteal revascularization (ΔABI = 0.05; ΔPACT-DMS = 2), whereas Patient 20L underwent femoral, infrapopliteal and pedal-vessel revascularization (ΔABI = 0.11; ΔPACT-DMS = 5). Postoperative images showed increased vessel filling, clearer visualization of previously weak vascular structures and increased small-vessel signals. The broader changes in Patient 20L were qualitatively consistent with the greater distal extent of intervention. Orange arrows indicate increased vessel filling, and purple arrows indicate improved small-vessel visibility. ΔABI denotes postoperative minus preoperative ABI, whereas ΔPACT-DMS denotes the number of imaged toes showing visible postoperative improvement in PACT-DMS-derived vascular phenotypes relative to baseline. Grayscale intensity represents relative PA signal amplitude. L and R indicate the left and right feet, respectively. Scale bar, 2 mm. Clinical and ABI data are provided in Supplementary Tables 2 and 3; additional paired comparisons are shown in Supplementary Figs. 15–17.

Together, these exploratory observations suggest that preoperative PACT-DMS may provide an interpretable representation of distal vascular-bed status, whereas PACT-DMS-derived postoperative phenotypes may depict distal vascular refilling and improved small-vessel visibility after revascularization. This perioperative analysis remains exploratory and should not be interpreted as a validated prognostic or treatment-response model.

## Discussion

A central challenge in PAD assessment is that restoration or preservation of large-vessel patency does not necessarily ensure adequate distal tissue perfusion. Symptom relief, wound healing and long-term limb outcomes depend substantially on the structure and function of the distal vascular bed, yet current non-invasive tools provide limited structurally resolved information at this scale. In this study, we developed a clinic-oriented multispectral compound-scanning photoacoustic tomography system and an interpretable PACT-based distal vascular phenotyping framework, termed PACT-DMS, to quantify PAD-related distal vascular status from seven physiologically motivated vascular features. In a prospective single-centre cohort, PACT-DMS showed promising discrimination between patients with PAD and healthy controls. Exploratory analyses further suggested that the framework may provide complementary information in clinically diagnosed PAD limbs with non-abnormal ABI and may support perioperative assessment of distal vascular-bed reserve and postoperative vascular refilling. These findings support MCPATS-enabled photoacoustic distal vascular phenotyping as a candidate adjunctive approach for tissue-level vascular assessment in PAD, rather than as a replacement for established vascular tests.

The biological interpretation of PACT-DMS is that PAD-related distal vascular impairment is not simply a reduction in trunk-vessel calibre. The seven features captured coordinated changes in trunk-vessel morphology, small-vessel clustering or redistribution, topological coupling between larger vessels and small-vessel networks, and pulsation-related dynamic behaviour. This multidimensional phenotype is consistent with the broader view that PAD involves both large-vessel obstruction and distal small-vessel dysfunction, and that limb status is shaped by tissue-level perfusion rather than proximal arterial patency alone[10, 26]. By resolving these structural and dynamic phenotypes, PACT-DMS provides an imaging-based representation of PAD as a disorder of the terminal perfusion bed, linking macroscopic arterial disease to distal vascular-bed remodelling.

PACT-DMS extends prior photoacoustic and optoacoustic work in three concrete ways. First, whereas raster-scan optoacoustic mesoscopy has been used to score diabetic skin microangiopathy from dermal morphophysiological features[61, 62], PACT-DMS targets the foot trunk-and-small-vessel network directly relevant to PAD-related distal perfusion and uses physiologically labelled L/T/S vessel categories rather than skin-layer features. Second, whereas radiomic analysis of photoacoustic foot images has been explored for predicting perfusion-related status[60], PACT-DMS uses a low-dimensional, clinically readable feature set that remains traceable to prespecified vascular phenotypes. Third, compared with tests that provide indirect tissue oxygenation or superficial vascular readouts, including TcPO₂, skin perfusion pressure, hyperspectral imaging, OCT angiography, contrast-enhanced ultrasound and nailfold capillaroscopy, PACT-DMS provides contrast-free and compression-free structural – dynamic phenotyping of the foot-pulp vascular bed over a centimetre-scale field of view with submillimetre spatial resolution. PACT-DMS therefore occupies a distinct niche as a contrast-free, compression-free photoacoustic readout of foot-pulp vascular-bed structure and dynamics with feature-level interpretability.

Clinically, the potential value of PACT-DMS lies in its ability to add a distal vascular-bed readout to current PAD assessment pathways. ABI, ultrasound, CTA and other routine tests remain essential for haemodynamic screening, anatomical localization and treatment planning[8], but they do not directly characterize the morphology, distribution and dynamic behaviour of the PACT-resolved distal vascular bed. In clinically diagnosed PAD limbs with non-abnormal ABI, PACT-DMS identified abnormal distal vascular phenotypes, suggesting that photoacoustic phenotyping may provide information not fully reflected by pressure-based indices. Similarly, perioperative analyses suggested that preoperative PACT-DMS may reflect distal vascular-bed reserve, whereas PACT-DMS-derived postoperative phenotypes may visualize distal vascular refilling and small-vessel visibility after revascularization. These observations should be interpreted as exploratory, but they indicate a plausible clinical niche for PACT-DMS as a complementary imaging readout when macrovascular indices and tissue-level vascular status are not fully concordant. A methodological strength of this study is the transformation of high-resolution PACT images into a low-dimensional, interpretable feature space, rather than reliance on high-dimensional radiomic signatures or complex deep-learning models. The use of semantic vascular features and a linear SVM makes the decision score traceable to concrete vascular phenotypes, which may facilitate clinical interpretation, cross-study comparison and future standardization. The representative-section strategy was not intended to replace complete three-dimensional modelling, but to provide a stable and anatomically constrained representation suitable for a small clinical cohort; adjacent-section consistency supported this approach, although future studies should compare representative sections, multi-section aggregation and full-volume feature modelling. From a translational perspective, because PACT-DMS relies on a limited number of interpretable structural and dynamic features rather than fully validated multispectral oxygenation quantification, its analytical concept may be adaptable to simplified implementations such as single-wavelength two-dimensional tomography or limited-range serial-section scanning.

Exploratory multispectral observations suggested that local vascular signal heterogeneity may contain additional biological information beyond the seven-feature PACT-DMS framework. In Supplementary Fig. 18, small-vessel spectra showed oxygenated-haemoglobin-like patterns in a healthy volunteer but more deoxygenated-haemoglobin-like features in a patient with PAD; reduced large-vessel signal intensity and heterogeneous, flocculent intravascular patterns were also observed in patients with poor postoperative recovery. These observations suggest potential future directions for studying distal vascular oxygenation, vascular contents and local tissue microenvironment, but should be regarded as hypothesis-generating because they were based on few cases and lacked independent biochemical, pathological or haemodynamic validation. These findings should be interpreted within the context of several methodological and translational limitations. First, this was a single-centre study with 24 patients with PAD and 21 healthy controls, and subject-level leave-one-out estimates may still be optimistic in a small cohort; external validation across independent devices, operators and cohorts is required before clinical claims can be made. Second, healthy controls were free of diabetes by design, producing a near-complete diabetes imbalance between groups. Diabetes-stratified sensitivity analysis showed directionally consistent reductions in bilateral PACT-DMS scores in both diabetic and non-diabetic PAD subgroups relative to healthy controls, suggesting that the observed imaging differences were not solely dependent on diabetes distribution. However, this remains a sensitivity analysis rather than definitive confounding control, and larger cohorts with balanced diabetes prevalence are needed. Third, we did not perform systematic head-to-head comparisons with toe-brachial index, transcutaneous oxygen pressure, skin perfusion pressure or other established distal perfusion and microcirculatory tests; therefore, the incremental information of PACT-DMS beyond these methods remains unquantified. Fourth, the ABI-non-abnormal subgroup and perioperative analyses were small and exploratory, and should not be interpreted as definitive evidence of screening, prognostic or treatment-response utility. Fifth, the current workflow still relies on semi-automatic segmentation and observer-guided pulsation grading, both of which require further automation and standardization before multicentre deployment. At the present stage, multicentre validation is also practically constrained by the limited availability of standardized PACT systems, the need for cross-device calibration, harmonized acquisition and quality-control procedures, operator training, and balanced recruitment of PAD and control participants with comparable diabetes status and disease severity. Future multicentre studies should therefore use locked feature definitions, prespecified classifier settings, standardized acquisition workflows and inter-system reproducibility testing to define the diagnostic performance, clinical application boundaries and translational feasibility of PACT-DMS.

## METHODS

### MCPATS system for compression-free distal foot photoacoustic imaging

Distal foot vascular structure, multispectral signals and dynamic PACT images were acquired using MCPATS, a custom-built multispectral compound-scanning photoacoustic tomography platform. The MCPATS implementation combined an open curved-array imaging probe, temperature-controlled water coupling, foot stabilization, position-memory local rescanning, multispectral excitation and energy-normalized data acquisition. The system supported rapid three-dimensional toe imaging, repeatable local rescanning at predefined anatomical positions, and multispectral or dynamic sequence acquisition. In compound-scanning mode, multi-angle acquisition was used to improve three-dimensional vascular visualization and reduce directional sampling dependence, whereas rapid single-angle scanning was used when acquisition efficiency was prioritized. Foot-shape-informed scanning-path planning was used to support individualized coverage of the distal foot and toes.

The imaging probe used a 5.5 MHz centre-frequency ultrasound transducer array with 60% bandwidth, comprising 256 elements uniformly distributed along a 180° arc with an 80 mm radius. Each element used a self-focusing design with a focal depth of 70 mm, providing a uniform imaging field of view of approximately 2 cm in radius. The open curved-array geometry and water-coupling configuration enabled toe imaging without direct mechanical compression, which is important for superficial distal vessels that are small and sensitive to external pressure.

Photoacoustic excitation was provided by a dedicated optical parametric oscillator based on type-I BBO crystal phase matching and a short resonant cavity, with optical energy delivered through an optical fibre bundle. The OPO covered the 700–950 nm wavelength range and generated a skin-surface illumination region of approximately 4 cm × 0.5 cm, with a fluence of approximately 10 mJ cm−2, below ANSI safety limits. Integrated energy and wavelength monitoring were used for frame normalization and spectral quality control. Photoacoustic signals were acquired using a custom-built data-acquisition card with shielding and grounding optimization, and the receiving impedance was set to 50 Ω to match the front-end circuit and reduce electrical noise and signal tailing. For compactness and clinical acquisition stability, the system incorporated a folded optical path, an independently ducted water-cooling module with elastic support, and air-spring vibration isolation of the optical module. The MCPATS configuration, OPO design and spatial-resolution characterization are shown in Supplementary Figs. 1–3.

### Imaging protocol and clinical testing procedure

This was a single-centre, prospective, observational diagnostic study. We recruited 24 patients with clinically and radiologically confirmed PAD and 21 age-matched healthy volunteers. Healthy controls were selected to reduce major vascular and metabolic confounding: they had no clinical diagnosis of PAD, no history of diabetes, no stage 5 chronic kidney disease or uraemia, and no severe foot trauma. Smoking and alcohol consumption were not used as exclusion criteria to preserve control-sample representativeness. By design, healthy controls were free of diabetes, end-stage renal disease and severe foot trauma to reduce metabolic and structural confounding. This produces a near-complete diabetes imbalance between groups (PAD 70.8% versus controls 0.0%), which we address through diabetes-stratified sensitivity analysis (Supplementary Fig. 13b) and discuss as a limitation. A confirmatory study should enrol non-diabetic PAD versus non-diabetic controls in matched proportions.

The study protocol was approved by the Ethics Committee of Beijing Friendship Hospital, Capital Medical University (approval number 2024-P2-080-01), and all participants provided written informed consent before examination.

To minimize physiological and environmental influences on peripheral vascular status, imaging was performed under standardized conditions. Participants avoided strenuous exercise for at least 1 h before examination and rested for more than 10 min in a room maintained at 20–30 °C. Imaging was performed in the seated position with standardized chair height, foot stabilization and avoidance of lower-limb compression by clothing or posture. The coupling-water temperature was maintained at 30–35 °C; quality-control analysis showed that mild variations within this range did not cause systematic changes in visible vascular morphology (Supplementary Fig. 4). Coupling gel, disposable sterile water-tank liners, probe disinfection and laser-safety eyewear were used to reduce artefacts and ensure examination safety.

Each toe of both feet underwent rapid three-dimensional PACT volume scanning with a step size of 0.1 mm. Key regions were then selected from real-time three-dimensional reconstructions and cross-sectional images for local multispectral acquisition when required. The diagnostic PACT-DMS model used the representative section of the second toe, whereas postoperative exploratory assessment used serial sections across multiple toes.

### Image reconstruction and display

Raw photoacoustic signals were band-pass filtered at 1–10 MHz to suppress low-frequency drift and out-of-band electrical noise. Before reconstruction, angular Fourier interpolation was applied to increase the 256-channel array data to 512 angular sampling points and reduce undersampling-related streak artefacts. Images were reconstructed using a delay-and-sum algorithm without post-reconstruction filtering or negative-value removal. Reconstruction was implemented with CUDA-based parallel computation, enabling real-time cross-sectional imaging and three-dimensional toe-volume visualization by maximum-intensity projection. All images used for quantitative analysis were reconstructed with fixed parameters, fixed pixel size and a unified intensity-normalization workflow; display adjustments were used only for real-time localization and were not involved in feature calculation.

For three-dimensional volume analysis, SAM-assisted tissue-boundary segmentation was used to suppress regions outside the tissue boundary and deep non-target regions more than 1 cm from the surface, reducing skin signal, reflection artefacts and undersampling-related background interference. Vessel images were analysed and displayed in grayscale to emphasize vascular morphology and avoid potential bias from inconsistent pseudocolour rendering.

### Dataset and preprocessing

Three-dimensional PACT data were acquired from all five toes of both feet in each participant. The second toe was selected as the primary analysis region because it showed relatively consistent vascular orientation and lower morphological variability across participants, facilitating anatomically comparable feature extraction. The anatomical rationale and PCA-assisted representative-section selection workflow are shown in Supplementary Fig. 7. After reconstruction of the second-toe volume, preprocessing included vessel enhancement, image segmentation and intensity normalization. Volumes with insufficient signal-to-noise ratio or reconstruction quality, as well as invalid limbs from participants with unilateral disability, were excluded. This yielded 44 PAD limbs and 40 healthy limbs for limb-level analysis. Limb status was determined by integrated assessment of clinical diagnosis, symptomatic side and vascular imaging data, including CTA or digital subtraction angiography. Because both lower limbs of the enrolled patients with PAD showed varying degrees of disease-related vascular involvement, both feet were included as PAD limb-level observations. Model validation was performed using subject-level leave-one-out cross-validation to avoid bilateral information leakage. De-identified clinical and vascular imaging summaries are provided in Supplementary Data 2.

To obtain a standardized low-dimensional analysis section without using diagnostic labels, PCA-assisted representative-section selection was performed within a predefined anatomical window in the proximal phalanx segment of the second toe. Let the reconstructed PACT volume in this region be represented by a sequence of slices:

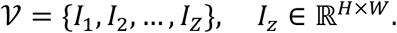

The vectorized form of slice *I_z_* was denoted by *x_z_*:

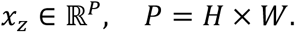

The slice vectors were concatenated to form the Casorati matrix

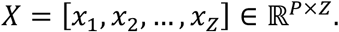

The column mean was calculated as

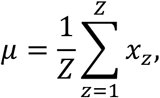

and the mean-centred matrix was written as

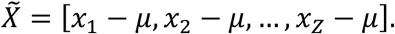

Singular-value decomposition was then performed:

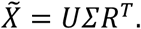

The first right singular vector *r*_1_ describes the dominant depth-dependent vascular structural pattern within the predefined region. The representative slice was selected as

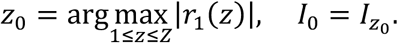

Because this rule was applied within a prespecified anatomical region and did not use disease labels, classification outcomes or visually selected lesions, it was intended to reduce operator-preference-driven section selection while preserving a standardized, anatomically constrained representation for interpretable feature extraction. Adjacent-section consistency analysis is shown in Supplementary Fig. 8.

Within each representative section, visible vascular structures were assigned to three categories: longitudinal vessels (L-vessels), transverse vessels (T-vessels) and small vessels (S-vessels). L-vessels run along the toe axis and represent major distal perfusion conduits; T-vessels are oblique or transverse branches extending from longitudinal vessels; and S-vessels include discrete fine vessels or locally aggregated small-vessel networks. Based on this classification, seven predefined PACT-DMS features were extracted: L-vessel area, L-vessel circularity, vessel pulsation strength, Clustered S-vessel area, T–L vessel-count difference, S-vessel count and S-T/L intersection count. These features quantified trunk-vessel morphology, perfusion-related dynamic behaviour, microvascular clustering, vessel-type redistribution and trunk-microvascular coupling.

Arteries and veins were not separately labelled during PACT-DMS feature construction. At the distal toe level, stable arteriovenous discrimination based solely on multispectral signatures was not always feasible under the present imaging conditions, even when superficial arteries and veins could be morphologically distinguished (Supplementary Fig. 19). In addition, the aim of PACT-DMS was to phenotype the terminal vascular bed as an integrated structure-function network rather than to assign each small vessel to an arterial or venous identity. Therefore, all visible vascular structures were analysed within the L/T/S morphological framework.

### Feature extraction and diagnostic-model development

After representative-section selection, visible vascular structures were segmented and assigned to the L/T/S morphological framework. Two photoacoustic imaging researchers with clinical backgrounds and more than 2 years of experience independently delineated vessel boundaries in 3D Slicer after standardized training and pre-annotation using non-feature sections. Discrepant segmentations were reviewed by consensus; unresolved cases were adjudicated by a third photoacoustic imaging researcher with more than 5 years of experience. Image segmentation, L/T/S annotation and pulsation assessment were performed on de-identified PACT images without access to ABI values, postoperative outcomes or model outputs; details of blinding and independent review are provided in Supplementary Note 3. Interobserver agreement was quantified using foreground-only Dice coefficients to avoid overestimation by large background regions. The median Dice coefficient was 0.70 (IQR, 0.64–0.75), with 51.9% of cases having Dice ≥ 0.60, indicating moderate interobserver consistency for small-vessel foreground segmentation under the present semi-automatic annotation workflow. Segmentation consistency and representative examples are shown in Supplementary Fig. 11. Let the segmented vascular label map on the representative section be

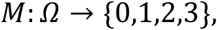

where *Ω* is the image-plane pixel set; 0 denotes background, 1 longitudinal vessels (L-vessels), 2 transverse vessels (T-vessels) and 3 small vessels (S-vessels). The three vascular subregions were defined as

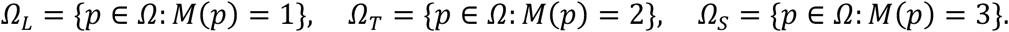

Connected components within each subregion were denoted by *C_L_*, *C_T_* and *C_S_*, and their component counts by *N_L_*, *N_T_* and *N_S_*. Based on these labelled regions, seven predefined semantic PACT-DMS features were extracted to quantify trunk-vessel morphology, perfusion-related dynamic behaviour, microvascular clustering, vessel-type redistribution and trunk–microvascular coupling.

F1, L-vessel area, was defined as the maximum physical area among L-vessel connected components:

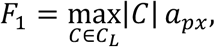

where *a_px_* is the physical area of a pixel.

F2, L-vessel circularity, described the morphological irregularity of the largest L-vessel component. For an L-vessel component with area *A* and perimeter *P*, classical circularity was first calculated as 4*πA*/*P*^2^, and the feature was defined as a logarithmic inverse-circularity score:

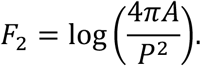

A more circular trunk-vessel cross-section yields an F2 value closer to 0, whereas a flattened, elongated or irregular cross-section yields a more negative F2 value; therefore, lower F2 values indicate impaired trunk-vessel morphological integrity. For model construction, F2 was standardized with the other features within each training fold, and its signed contribution to PACT-DMS was determined by the learned coefficient of the linear SVM decision function, whose direction was oriented so that lower scores indicated more PAD-like phenotypes.

F3, vessel pulsation strength, was derived from serial PACT images rather than from a single representative section, and was not a direct measurement of blood-flow velocity or volume. Pulsation was first graded according to whether serial frames showed vessel displacement, morphological change or filling change with a clear pulse-like cyclic pattern. In the diagnostic model, this dynamic phenotype was operationalized as a binary indicator:

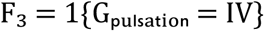

where grade IV indicated clear pulse-like cyclic vessel pulsation and grades I–III were encoded as 0. Two trained observers independently assessed pulsation; disagreements were resolved by consensus review and third-observer adjudication when necessary. Cohen’s kappa for binary pulsation assessment was 0.764 (95% CI, 0.628–0.901), with an observed agreement of 88.2%. Pulsation-scoring criteria and representative examples are shown in Supplementary Fig. 9, observer agreement is shown in Supplementary Fig. 10, and raw observer scores, adjudicated results and final binary pulsation labels are provided in Supplementary Data 5.

F4, Clustered S-vessel area, quantified local small-vessel aggregation. DBSCAN was applied to pixel coordinates in *Ω_S_*. If *K_S_* is the set of small-vessel clusters satisfying the minimum-sample threshold and *A_k_* is the area of the kth cluster, then

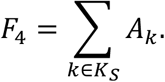

F5, T–L vessel-count difference, described vessel-type redistribution between transverse and longitudinal structures:

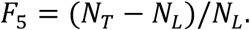

F6, S-vessel count, was the number of small-vessel connected components:

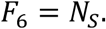

F7, S-T/L intersection count, quantified the coupling between small vessels and the trunk-vessel network. If *I_S_*_,*T*/*L*_ denotes the set of local small-vessel intersections with L– or T-vessels, then

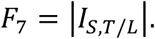

Examples of L/T/S vessel classification and the evidence basis for the seven PACT vascular features are shown in Supplementary Figs. 5 and 6, and feature definitions with potential pathophysiological interpretations are provided in Supplementary Table 5. The final feature vector for each limb-level sample was

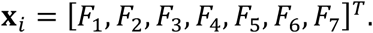

Given the low-dimensional, prespecified feature set and the study’s emphasis on clinical interpretability, a linear SVM was chosen over more complex nonlinear or ensemble classifiers so that each decision score remained directly traceable to the weighted contributions of the seven vascular features. Feature standardization was performed using training-fold parameters only and then applied to the held-out samples. Detailed model settings, including the SVM regularization parameter, class-weighting strategy, decision-threshold definition and fold-wise standardization procedure, are provided in Supplementary Note 3 and Supplementary Table 6; all preprocessing parameters used for standardization and thresholding were derived without access to the held-out participant. Let **z***_i_* be the standardized feature vector of the ith sample and *y_i_* ∈ {−1, +1} the class label. The linear SVM decision function was

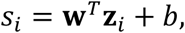

where **w** is the weight vector and *b* is the bias term. The model was trained by solving the soft-margin optimization problem

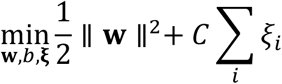

subject to

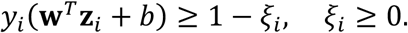

The signed decision score was used as the PACT-DMS score and was oriented so that lower values indicated a more PAD-like distal vascular phenotype and higher values indicated a more healthy-reference phenotype. Although the SVM decision score was oriented toward the healthy-reference phenotype, all clinical performance metrics were reported with PAD treated as the positive class.

Because PAD is systemic but bilateral involvement may be asymmetric, two analytical perspectives were retained. In unilateral analysis, each foot was treated as a limb-level observation to characterize side-specific distal vascular phenotypes, but not as an independent subject-level sample. In bilateral analysis, left– and right-foot SVM decision scores from the same participant were averaged to generate a participant-level fusion score:

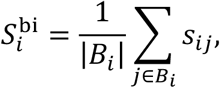

where *B_i_* denotes the available foot sides of participant *i*. When both sides were available, |*B_i_*| = 2; when only one valid side was available, fusion reduced to the unilateral score.

Model evaluation used subject-level leave-one-out cross-validation. In each fold, all available samples from one participant were held out as the test set, ensuring that bilateral feet from the same participant never appeared in both training and test sets. This prevented bilateral information leakage and better matched the clinical scenario of evaluating an unseen participant. All seven predefined features were retained in every fold, and no supervised feature selection or threshold tuning was performed within the test data. Model performance, permutation testing and leave-one-feature-out ablation results are shown in Supplementary Fig. 12; covariate sensitivity analyses are shown in Supplementary Fig. 13.

### Postoperative PACT-DMS-derived phenotypic assessment

For postoperative assessment, the complete seven-feature diagnostic PACT-DMS model was not reapplied as a treatment-efficacy score. Instead, paired serial sections across multiple toes were evaluated using two PACT-DMS-derived phenotypic dimensions linked to F2 and F6: changes in longitudinal-vessel morphology and fullness, and changes in small-vessel abundance and visibility, respectively. Early changes after revascularization were expected to reflect refilling of existing distal vessels and improved small-vessel visibility, rather than large-scale neovascular structural remodelling. For each toe, visible postoperative improvement was identified when enhanced trunk-vessel filling and/or increased small-vessel visibility was observed relative to baseline. ΔPACT-DMS was defined as the number of imaged toes showing such improvement, ranging from 0 to 5, with higher values indicating more extensive distal vascular-bed improvement. Scores of 0, 1–3 and 4–5 were interpreted as no visible improvement, limited improvement and marked improvement, respectively. This semiquantitative readout was distinct from the diagnostic linear-SVM PACT-DMS decision score. Multi-case paired examples illustrating postoperative changes in distal vessel filling, small-vessel visibility and local vascular-structure visualization are shown in Supplementary Figs. 15–17.

### Statistical analysis

Model performance was evaluated using accuracy, sensitivity, specificity, precision, F1 score, balanced accuracy and the area under the receiver operating characteristic curve (AUC). For these clinical performance metrics, PAD was treated as the positive class. For the exploratory ABI-related limb-level comparison, abnormal ABI was defined as ABI < 0.90, and non-abnormal ABI was defined as ABI ≥ 0.90. Implementation details of the classifier, thresholding strategy and fold-wise preprocessing are summarized in Supplementary Note 3 and Supplementary Table 6. In the unilateral analysis, metrics were calculated at the limb level; in the bilateral fusion analysis, metrics were calculated at the participant level after averaging the left– and right-foot SVM decision scores when both sides were available. AUC was calculated from receiver operating characteristic curves derived from continuous SVM decision scores. After thresholding the decision scores, classification metrics were defined as:

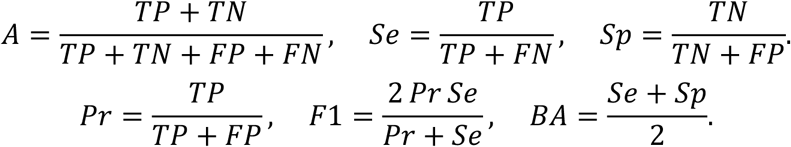

Here, *A*, *Se*, *Sp*, *Pr*, *BA*denote accuracy, sensitivity, specificity, precision and balanced accuracy, respectively. Confidence intervals for AUC and threshold-dependent classification metrics, including accuracy, sensitivity, specificity, precision, F1 score and balanced accuracy, were estimated using bootstrap resampling with 1,000 iterations. For unilateral analyses, resampling was performed at the participant level to preserve the pairing of bilateral limb measurements when both limbs were available. For bilateral fusion analyses, resampling was performed at the participant level. The 2.5th and 97.5th percentiles of the bootstrap distribution were reported as the 95% confidence interval. Permutation testing with 10,000 participant-level label permutations was used to assess whether the observed discrimination exceeded chance. Leave-one-feature-out ablation was performed by removing one predefined feature at a time and repeating the same subject-level cross-validation procedure. Exploratory participant-level covariate sensitivity analysis was performed using the bilateral fusion PACT-DMS decision score to assess its association with PAD status after adding age, sex and hypertension. Because model training was supervised, diagnostic labels were necessarily used during training; however, all fold-wise feature standardization, thresholding and model fitting were performed within the predefined subject-level cross-validation framework without access to the held-out participant. Detailed classifier implementation, threshold definition and leakage-control procedures are provided in Supplementary Note 3. These analyses were used to estimate uncertainty, assess chance-level separation, evaluate dependence on individual features and examine robustness to key participant-level covariates. For group-wise comparison of the seven PACT-DMS features, the Wilcoxon rank-sum test was used because of the limited sample size and non-Gaussian feature distributions. Benjamini-Hochberg false-discovery-rate correction was applied across the seven parallel feature tests, and *q* < 0.05 was considered statistically significant. Cliff’s delta was calculated as a non-parametric effect size:

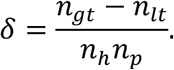

Here, *n_gt_* is the number of healthy-PAD feature-value pairs satisfying *x_ℎ_* > *x_ℎ_*, x_p_ is the number of pairs satisfying *x_ℎ_* < *x_ℎ_*, and *n*_ℎ_ and *n_p_* denote the healthy and PAD sample sizes, respectively. Clinical variables were compared using two-sided unpaired t-tests for approximately normally distributed variables and non-parametric tests for skewed variables. Exploratory participant-level covariate sensitivity analysis was performed to assess the association between bilateral fusion PACT-DMS decision score and PAD status after adding age, sex and hypertension; corresponding data are provided in Supplementary Data 1 and Supplementary Data 4, and results are shown in Supplementary Fig. 13.

Model performance, permutation testing and leave-one-feature-out results are shown in Supplementary Fig. 12, and covariate sensitivity analyses are shown in Supplementary Fig. 13. These analyses were used to quantify uncertainty, assess whether the observed discrimination exceeded chance, evaluate dependence on individual features and examine robustness to key participant-level covariates. Unless otherwise stated, all statistical tests were two-sided.

## Supporting information

Supplementary Information for Interpretable photoacoustic phenotyping of distal microcirculation for peripheral artery disease diagnosis with explorat

## Acknowledgments

We thank Chuhua Wu, Yan Luo, Yuwen Chen, Qianxi Wu of the Ma Lab for their support and helpful discussions. We are particularly grateful to Xiangzhi Su, Mingyue Yao and Zhe Zhao for their valuable discussions and suggestions during this study. We also thank Kun Wang and Zhenyu Liu from the Institute of Automation, Chinese Academy of Sciences, and Chao Zhang and Chenyi Guo from the intelligent healthcare research team at the Department of Electronic Engineering, Tsinghua University, for their insightful suggestions on the experimental design.

## Author Contributions

H.D., T.Y. and Z.L. contributed equally to this work. M.L and C.M. conceived and supervised the study, designed the overall research framework, coordinated the multidisciplinary collaboration, interpreted the results and took primary responsibility for manuscript preparation and revision. H.D. and T.Y. implemented the imaging and analysis workflow, performed data acquisition, image processing, feature extraction, statistical analysis, figure preparation and manuscript drafting. Z.L., W.W., Y.B. and J.M.G. contributed to the development of the PACT-DMS scoring framework by providing clinical and pathophysiological expertise on vascular anatomy, distal microcirculatory impairment and PAD-related disease mechanisms. M.L. contributed to patient recruitment, clinical diagnosis, vascular assessment, clinical data interpretation and clinical resource coordination. Y.B. contributed to clinical coordination and interpretation of relevant clinical information. J.X., N.Z., W.F. and X.W. contributed to imaging system production, maintenance, technical support, equipment operation and data acquisition. W.W., J.M.G. and M.L. provided clinical supervision. C.M., W.W., J.M.G. and M.L. supervised the study. All authors reviewed, edited and approved the final manuscript.

## Reporting summary

STARD 2015 and TRIPOD+AI reporting checklists are provided as Supplementary Data 6 and Supplementary Data 7 to support transparent reporting of the diagnostic-accuracy and model-development components of the study.

## Data availability

De-identified individual-level data, model outputs and reporting checklists are provided with the article as Supplementary Data. Supplementary Data 1 contains participant baseline characteristics; Supplementary Data 2 contains de-identified clinical presentations and vascular imaging summaries; Supplementary Data 3 contains limb-level subject-level LOOCV results, PACT-DMS decision scores and F1–F7 feature values; Supplementary Data 4 contains participant-level bilateral fusion scores and classification results; Supplementary Data 5 contains vessel pulsation scores, binary decisions, adjudication results and final pulsation labels; Supplementary Data 6 contains the STARD 2015 reporting checklist; and Supplementary Data 7 contains the TRIPOD+AI reporting checklist. Because raw PACT images and clinical imaging data may contain identifiable anatomical information or be subject to hospital ethics and data-management restrictions, complete raw imaging data are available from the corresponding authors upon reasonable request and after ethics and data-use approval.

## Code availability

The code developed for this study has been deposited on Figshare (DOI: 10.6084/m9.figshare.32186799). The repository includes the reconstruction code and the data-analysis/post-processing scripts supporting PACT-DMS feature extraction, subject-level leave-one-out cross-validation and statistical analyses. The code will be available to editors and peer reviewers for evaluation during peer review and will be made publicly available upon publication.

