## Supplementary Information for Interpretable photoacoustic phenotyping of distal microcirculation for peripheral artery disease diagnosis with explorat for "Interpretable photoacoustic phenotyping of distal microcirculation for peripheral artery disease diagnosis with exploratory perioperative assessment"

**Overview of Supplementary Information**

This Supplementary Information provides additional details on system construction, imaging workflow, feature design, model robustness and exploratory clinical analyses that support the main study. These materials are intended to substantiate the central conclusion of the manuscript: PACT-DMS can serve as an interpretable photoacoustic phenotyping framework for the distal microcirculatory status in peripheral artery disease (PAD). Importantly, the analyses of ABI-insensitive limbs, perioperative follow-up and multi-wavelength spectral observations are exploratory. They should therefore be interpreted as supportive and hypothesis-generating observations rather than independently validated diagnostic, prognostic or treatment-response models. The Supplementary Data also include STARD 2015 and TRIPOD+AI reporting checklists to document how the diagnostic-accuracy and model-development components of the study were reported.

**Supplementary Notes**

**Supplementary Note 1. MCPATS imaging platform, acquisition workflow and quality control**

The MCPATS platform was developed to meet the need for stable photoacoustic imaging of distal foot vessels. It integrates optical excitation, acoustic reception, mechanical scanning, water coupling and foot fixation, enabling two-dimensional tomographic imaging, three-dimensional volumetric acquisition and local multi-wavelength scanning within the same system. The platform was designed to obtain distal vascular images without directly compressing the toe tissue and to improve acquisition consistency across participants and repeated scans. Spatial resolution characterization, spectral and energy monitoring, and water-temperature quality-control experiments together provided the technical basis for stable imaging of small distal toe vessels. The MCPATS system configuration, optical excitation module, spatial resolution and acquisition quality control are shown in Supplementary Figs. 1–4.

**Supplementary Note 2. Anatomical standardization and construction of semantic vascular features**

The PACT-DMS framework is not based on high-dimensional black-box radiomic features. Instead, it uses predefined semantic vascular phenotypes with anatomical and physiological interpretability. To improve comparability across participants, the second toe was selected as the primary diagnostic analysis region. Within a predefined anatomical segment, an anatomically constrained and PCA-assisted representative cross-section selection procedure was used to reduce operator-dependent slice selection. L/T/S vessel categories were used to describe longitudinal trunk vessels, transverse or branch vessels and small-vessel plexus structures, respectively. The seven derived features characterize trunk calibre and morphology, small-vessel clustering and distribution, vascular network redistribution, topological coupling and dynamic vascular behaviour. Feature design was informed by clinical observations of PAD-related vascular lesions by vascular surgeons, analyses of PACT-visible structures by photoacoustic engineers, and conventional evidence from pathology, CTA, DSA and TBI. Thus, the feature system was designed to provide a physiologically interpretable representation of distal vascular remodelling in PAD, rather than a purely data-driven collection of image descriptors. L/T/S vessel classification, the clinical rationale of the seven features, representative cross-section selection, adjacent-slice consistency, vessel pulsation scoring criteria and agreement analysis for binary pulsation assessment are shown in Supplementary Figs. 5–10. Feature definitions and physiological interpretations are provided in Supplementary Table 5. Limb-level F1–F7 values and model inputs are provided in Supplementary Data 3, and pulsation scores, binary assessments and final pulsation labels are provided in Supplementary Data 5.

**Supplementary Note 3. Segmentation consistency, representative-section stability and model robustness**

Because the PACT-DMS workflow involves representative cross-section selection, vascular annotation and low-dimensional feature extraction, additional analyses were performed to assess the stability and reproducibility of these key steps. Adjacent-section analysis was used to test whether the PCA-assisted representative cross-section reflected a locally continuous vascular structural pattern rather than an isolated or manually selected single image plane. Interobserver segmentation consistency was assessed to evaluate the reproducibility of vascular annotation and downstream feature extraction. At the model level, bootstrap resampling, permutation testing, leave-one-feature-out ablation and covariate sensitivity analysis were used to examine the internal robustness of the discriminative results, together with analytical strategies that account for the non-independence of bilateral limb measurements.

These supplementary analyses do not replace external validation, but they provide internal support for the discriminative ability observed in this single-centre cohort and suggest that the results were not primarily driven by subjective slice selection, segmentation variability, random label assignment or a single unstable feature category. Representative-section stability, pulsation-assessment agreement, segmentation consistency, model robustness and covariate sensitivity analyses are shown in Supplementary Figs. 8, 10–13 and the related Supplementary Tables. Limb-level subject-level LOOCV predictions and feature values are provided in Supplementary Data 3, participant-level bilateral fusion scores in Supplementary Data 4 and raw vessel pulsation ratings in Supplementary Data 5.

**Classifier implementation and leakage control**

The diagnostic PACT-DMS classifier was implemented as a linear support vector machine using the seven predefined vascular features as input. The regularization parameter was fixed at C = 1 for all cross-validation folds, and class weighting was set to none. Feature standardization was performed within each training fold: the mean and standard deviation were estimated from the training participants only and then applied to the held-out participant. No feature-standardization parameter, model parameter or supervised decision threshold was estimated using the held-out participant.

The continuous SVM decision function was used as the PACT-DMS decision score. The score direction was oriented so that lower values indicated a more PAD-like distal vascular phenotype and higher values indicated a more healthy-reference phenotype. Although the SVM decision score was oriented toward the healthy-reference phenotype, all clinical performance metrics were reported with PAD treated as the positive class. Binary classification was performed using the default zero threshold of the SVM decision function. In the bilateral fusion analysis, left- and right-foot decision scores from the held-out participant were averaged before thresholding. No class label or feature value from the held-out participant was used during model fitting, standardization, parameter tuning or threshold definition.

**Blinding and independent review**
Clinical diagnosis, limb-status assignment and routine vascular imaging interpretation were performed independently of PACT-DMS feature extraction and model outputs. For image-based feature extraction, observers reviewed de-identified PACT images with only the anatomical information required for orientation. Clinical group, ABI values, CTA/DSA findings, postoperative outcomes and final model predictions were not used during vessel segmentation, L/T/S annotation or pulsation assessment. Discrepant segmentations and pulsation scores were resolved by consensus review and, when necessary, third-observer adjudication.

Supervised statistical modelling necessarily used PAD/control labels during training; therefore, the statistical analysis should not be considered fully blinded to diagnostic status. To reduce optimism and information leakage, all feature standardization, model fitting, thresholding and robustness analyses were performed within the predefined subject-level cross-validation framework, and no information from the held-out participant was used during training-fold preprocessing or model construction.

**Supplementary Note 4. Exploratory analyses of ABI-insensitive limbs and perioperative assessment**

The analyses of ABI-insensitive limbs and perioperative imaging were designed to explore whether distal PACT phenotypes provide information complementary to pressure-based macrovascular assessment. In the ABI-related analysis, ABI results and PACT-DMS classifications were compared in limbs with clinically confirmed PAD, with particular attention to limbs with normal ABI but abnormal distal vascular phenotypes by PACT-DMS. These findings should not be interpreted as evidence that PACT-DMS can replace ABI or as definitive proof of diagnostic superiority over ABI. A more appropriate interpretation is that distal photoacoustic microcirculatory phenotypes may capture PAD-related vascular involvement that is not fully reflected by pressure-based indices. The perioperative analysis was also exploratory. Preoperative PACT-DMS phenotypes were used to examine their potential relationship with clinical recovery trajectories after revascularization, and serial postoperative PACT images were used to visualize changes in distal vascular filling, small-vessel visibility and local vascular phenotypes. These analyses were intended to generate clinically interpretable hypotheses about distal vascular reserve and tissue-level recovery after revascularization, rather than to establish a validated prognostic model or treatment-response tool. ABI and PACT-DMS classification comparisons, individual clinical follow-up information and pre- versus postoperative ABI values are provided in Supplementary Tables 2–4. Perioperative cases and postoperative changes in distal vascular phenotypes are shown in Supplementary Figs. 14–17. Individual-level de-identified clinical and vascular imaging summaries are provided in Supplementary Data 2, limb-level PACT-DMS classifications in Supplementary Data 3 and subject-level bilateral fusion results in Supplementary Data 4.

**Supplementary Note 5. Exploratory multi-wavelength spectral observations and considerations for arteriovenous analysis**

Although the PACT-DMS framework in the main manuscript primarily uses structural and dynamic vascular phenotypes, the MCPATS platform also supports multi-wavelength spectral acquisition. This Supplementary Information therefore includes exploratory multispectral observations to illustrate local vascular signal heterogeneity, potential oxygenation-related changes and imaging phenomena related to intravascular compositional differences in selected cases. These findings are currently hypothesis-generating and should not be interpreted as validated biochemical or prognostic biomarkers. In the distal toe vascular bed, stable artery-vein classification at the single-vessel level based solely on multi-wavelength linear unmixing remains challenging, because superficial veins, small arteries and microvessels may show overlapping spectral behaviour under the imaging conditions used in this study. Accordingly, the PACT-DMS analysis in the main manuscript treats the distal toe vascular bed as an integrated structural-dynamic phenotype and does not require stable arteriovenous classification of individual vessels. Future studies should combine standardized multi-wavelength acquisition, validated oxygenation quantification models and independent haemodynamic reference standards to clarify the physiological specificity of these spectral observations. Exploratory multi-wavelength spectral observations and examples of the difficulty of distinguishing arteries from veins in the distal toe vasculature are shown in Supplementary Figs. 18 and 19.

A Supplementary Data workbook provides de-identified baseline, clinical and vascular-imaging, LOOCV, bilateral-fusion, and vessel-pulsation data. The Notes worksheet documents de-identification and coding. Participant and case IDs are randomly assigned study codes; they have no intrinsic clinical or chronological meaning and do not reflect admission, recruitment, imaging, treatment, or follow-up order. Supplementary Data 1–5 provide traceable sources for the main text, figures and tables.

### Supplementary Figures

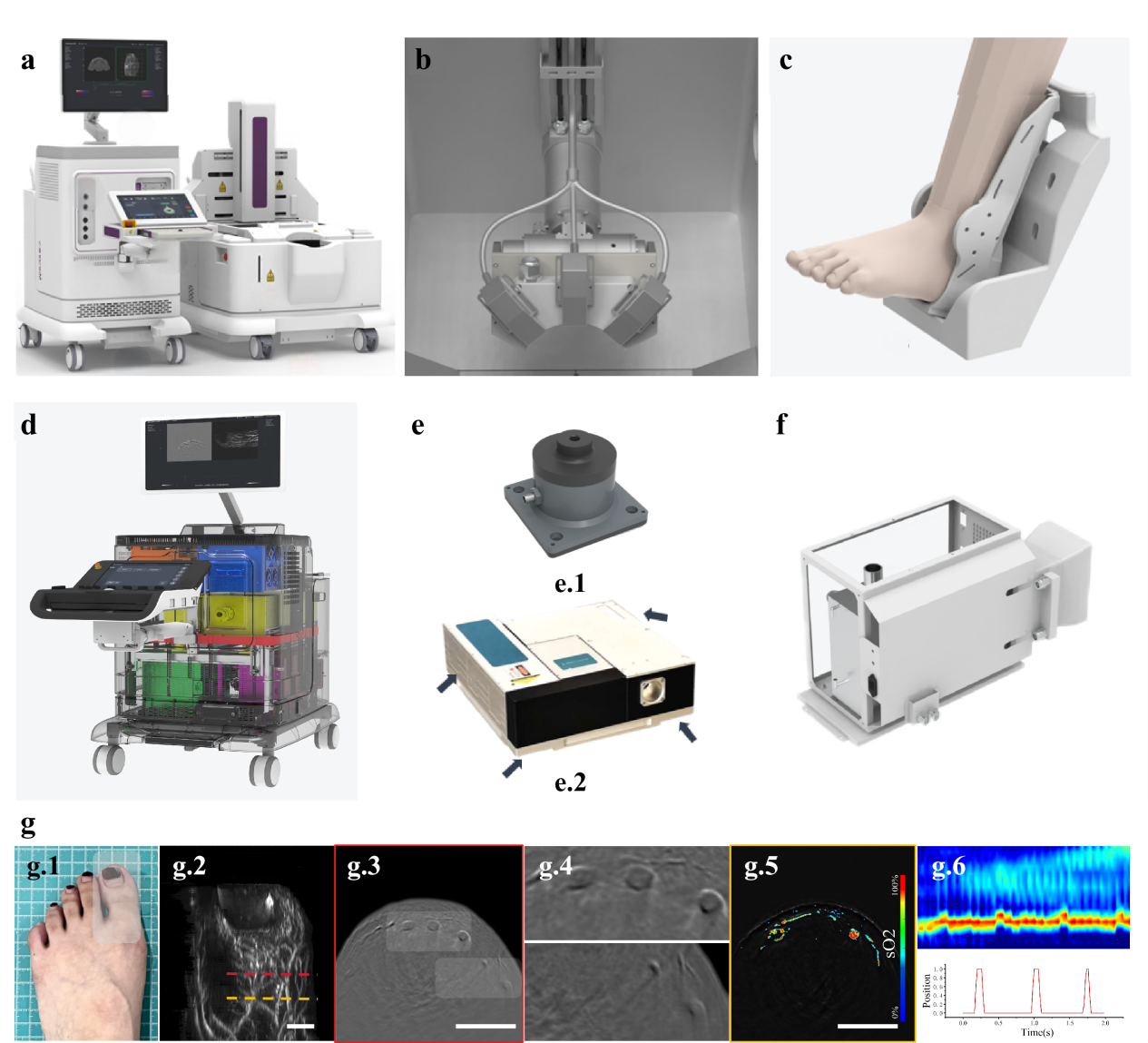

**Supplementary Fig. 1 | Architecture and imaging capabilities of the MCPATS system. a,** Overall view of the MCPATS, comprising an integrated console and a water-coupled scanning unit for optical excitation, acoustic detection, mechanical scanning, data acquisition and system control. **b,** Photoacoustic scanning head integrating the imaging probe, fibre-bundle illumination and motorized scanning components. **c,** Foot fixation device for stabilizing foot position and improving repeatability. **d,** Perspective view showing the integrated optical, acoustic, mechanical and control modules. **e,** Optical excitation module, including the air-spring vibration-isolation support (**e.1**) and optical parametric oscillator (**e.2**). **f,** Laser water-cooling and thermal-management module for stable prolonged operation. **g,** Representative information acquired by MCPATS, including the imaging site (**g.1**), three-dimensional vascular imaging (**g.2**), reconstructed tomographic images (**g.3, g.4**), oxygenation-related spectral information (**g.5**) and dynamic vascular signals (**g.6**). Together, these components support stable three-dimensional, multispectral and dynamic photoacoustic imaging of the distal foot vasculature.

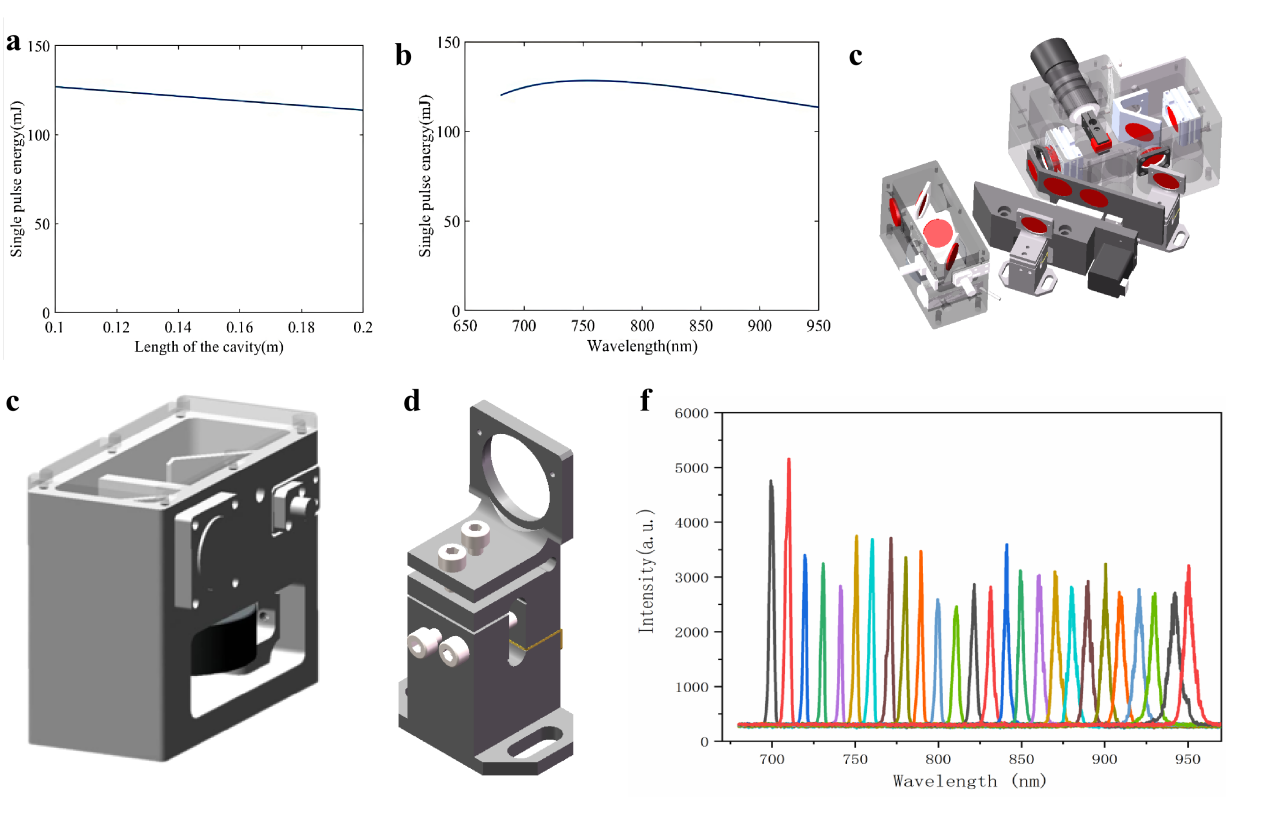

**Supplementary Fig. 2 |** **Theoretical design and engineering implementation of the optical parametric oscillator. a,** Relationship between output single-pulse energy and resonator cavity length calculated using coupled-wave equations, used to determine the cavity-length parameters needed for foot PACT imaging. **b,** Relationship between output single-pulse energy and wavelength, characterizing the energy distribution of the OPO within the target operating band. **c,** Engineering structure of the OPO head, showing compact integration of resonator mirrors, pump-beam coupling mirror, BBO crystal and galvanometer motor. **d,** Energy-monitoring module, used for real-time monitoring of output optical energy and frame-to-frame energy normalization. **e,** Details of the mirror mount, designed to improve optical-alignment precision and long-term mechanical stability. **f,** Output spectrum, characterizing the actual output within the target wavelength range. This excitation module supports multi-wavelength acquisition and energy/spectral monitoring, providing the engineering basis for subsequent exploratory spectral analysis and standardized image acquisition.

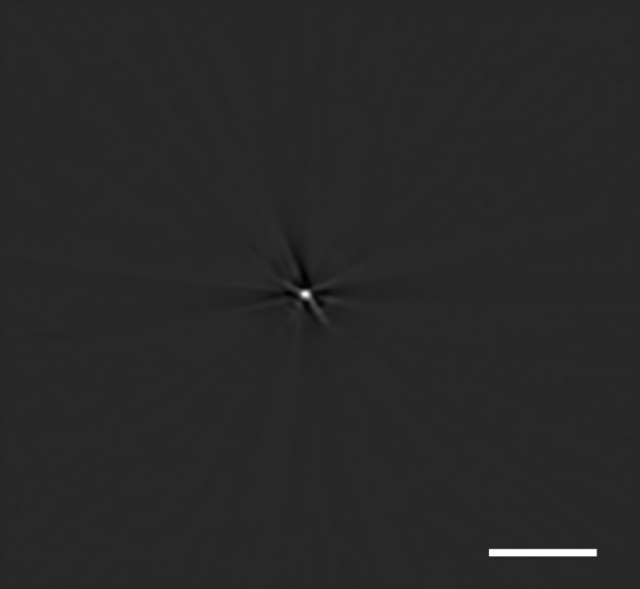

**Supplementary** Fig. 3 | Spatial resolution characterization of the system. A hair was used as an approximate line absorber for spatial-resolution testing. The hair was placed in water and adjusted into the imaging field of view. After PACT reconstruction, a normalized intensity profile was extracted along the transverse direction, and the full width at half maximum was used to characterize the effective spatial resolution. Because the hair has a finite diameter, the image profile reflects the effective response resulting from convolution of the system point-spread function and the target size. After subtracting an approximate target diameter of 50 μm, the spatial resolution of the system was estimated to be approximately 145 μm. This resolution supports visualization and morphological analysis of distal toe trunk vessels, small-vessel structures and their shape changes. Scale bar, 5 mm.

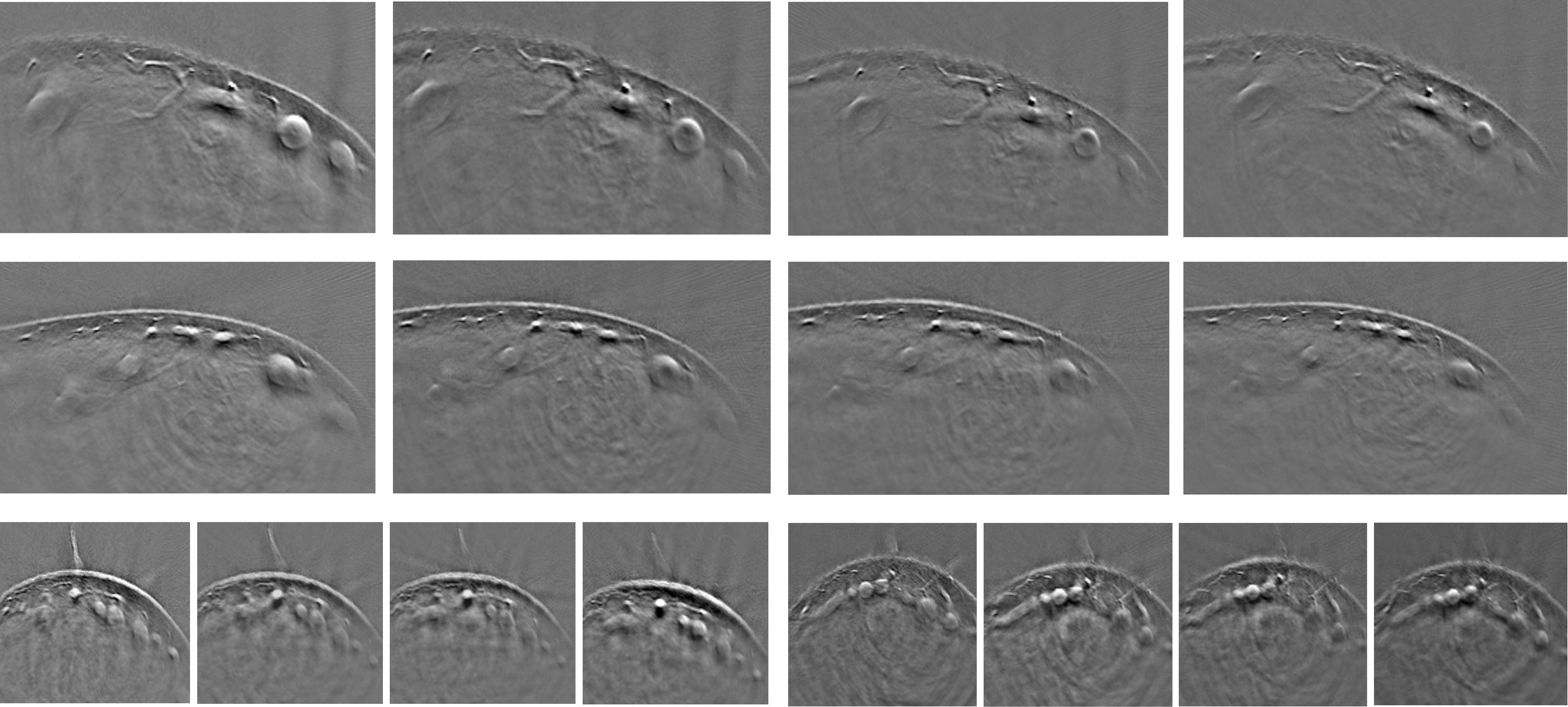

**Supplementary Fig. 4 | Effect of water temperature on visible vascular morphology.** The same volunteer underwent toe PACT imaging at different water temperatures: 40.0 °C, 33.5 °C, 30.0 °C and 26.5 °C. Representative cross-sectional images from multiple locations are shown. Within the vascular scale observable by this system and the water-temperature range used in the study, the visible trunk-vessel morphology did not show systematic changes. These results support the use of a standardized water-coupling temperature range of 30–35 °C in the main imaging workflow to reduce the effect of environmental conditions on interpretation of distal vascular morphology.

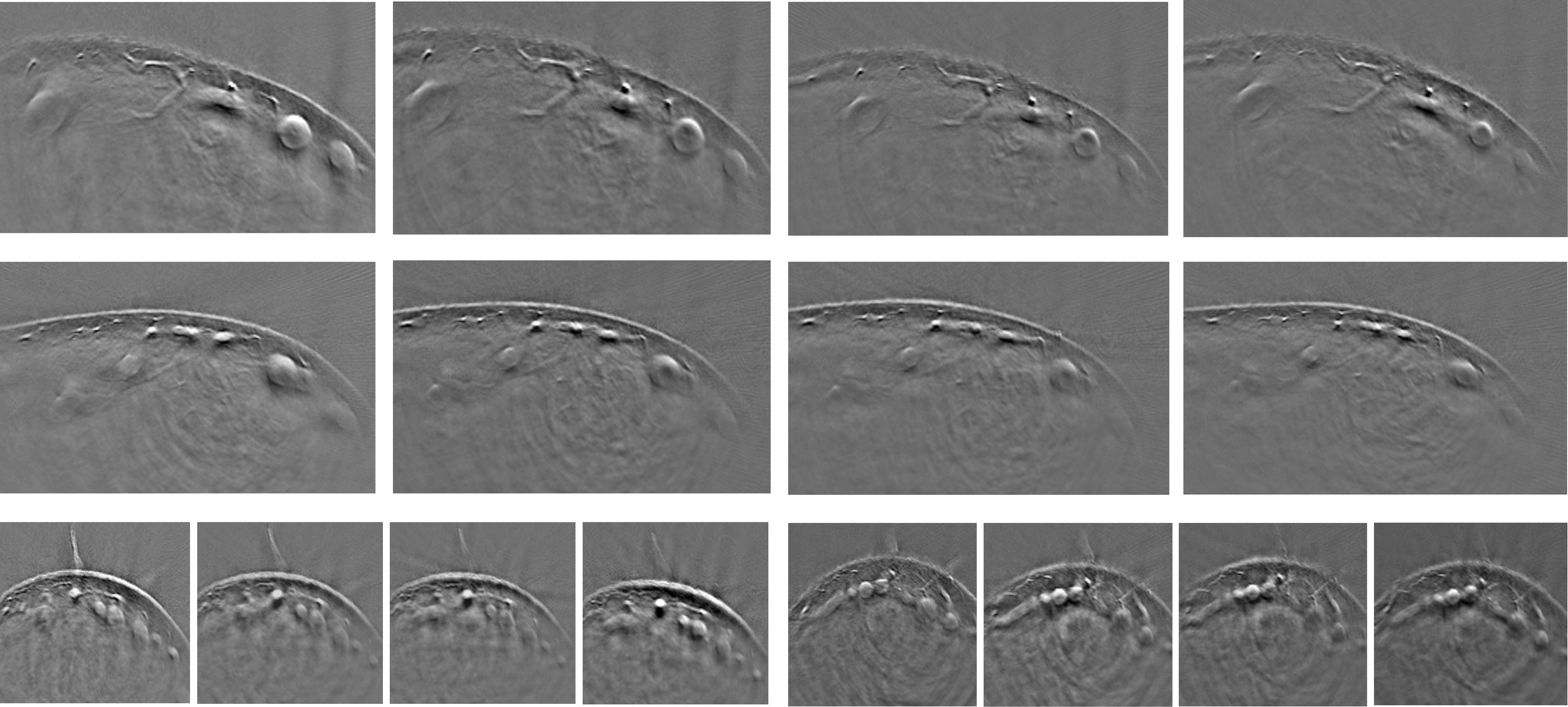

**Supplementary Fig. 5 | Examples of L/T/S vessel classification. Representative annotations of three types of vascular structures in PACT cross-sections are shown.** L-type vessels, marked in green, are longitudinal trunk vessels running approximately parallel to the long axis of the toe and can be considered the main structural pathway for distal perfusion. T-type vessels, marked in blue, are branches or cross-sectional connecting pathways extending obliquely or transversely from the longitudinal trunks. S-type vessels, marked in red, are small-vessel plexuses composed of finer branches with local clustered or reticular distributions. This classification converts complex distal vascular structures in PACT images into interpretable semantic units and provides the morphological basis for the seven vascular features used in the main manuscript.

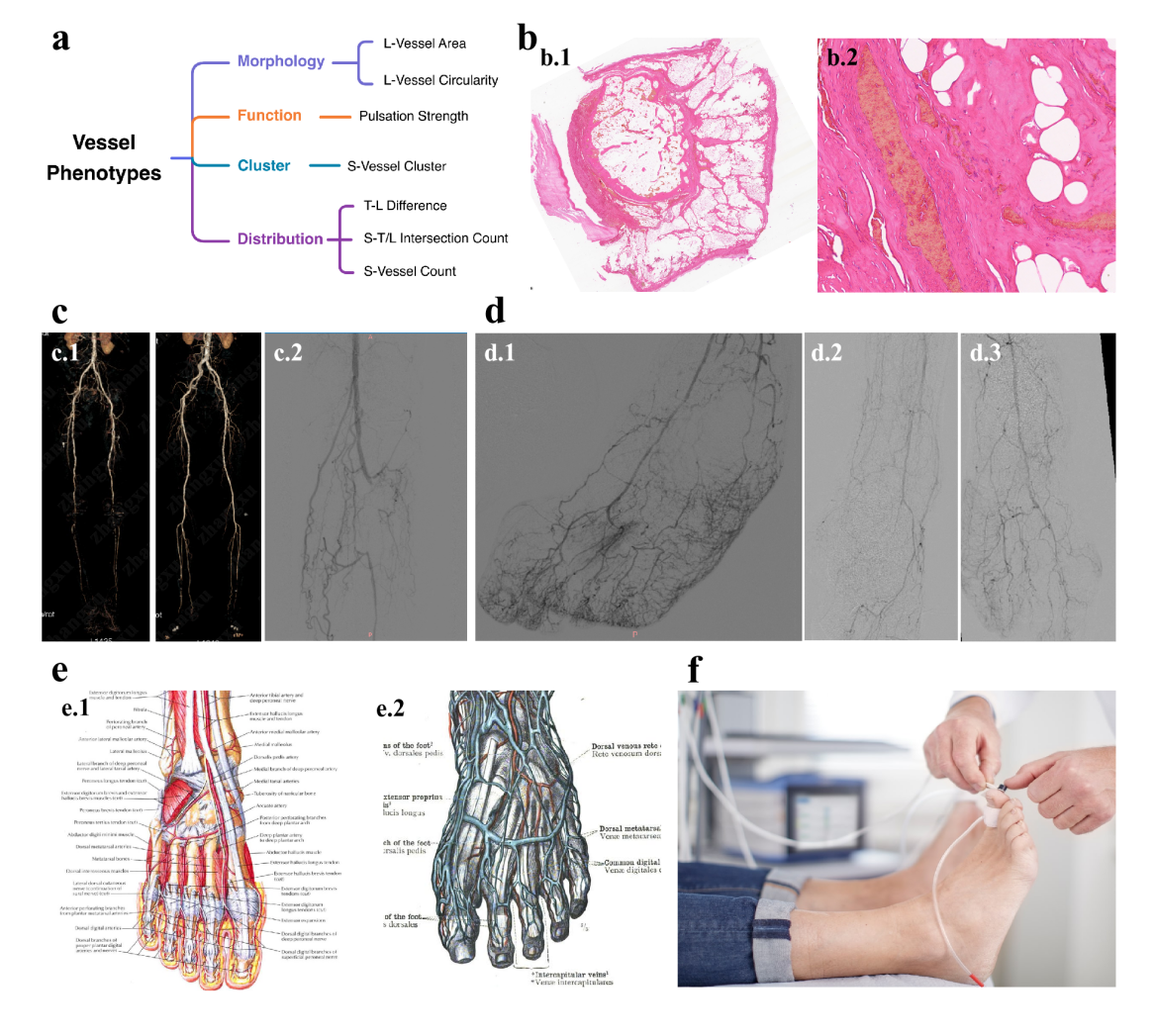

**Supplementary Fig. 6 | Clinical and anatomical basis of the seven PACT-DMS features. a,** Seven vascular features grouped into morphology, function, clustering and distribution. **b.1, b.2,** Toe histology showing organized thrombus and vascular remodelling. **c.1, c.2,** Lower-extremity angiography showing occlusion, collateralization and incomplete recanalization. **d.1–d.3,** Pedal angiography showing irregular distal vessels, network loss and reduced terminal branching. **e.1, e.2,** Normal pedal arterial and venous anatomy [1,2]. **f,** Toe–brachial index measurement illustrating toe-level haemodynamics. Together, these pathological, angiographic and anatomical references support the physiological interpretability of the seven PACT-DMS features.

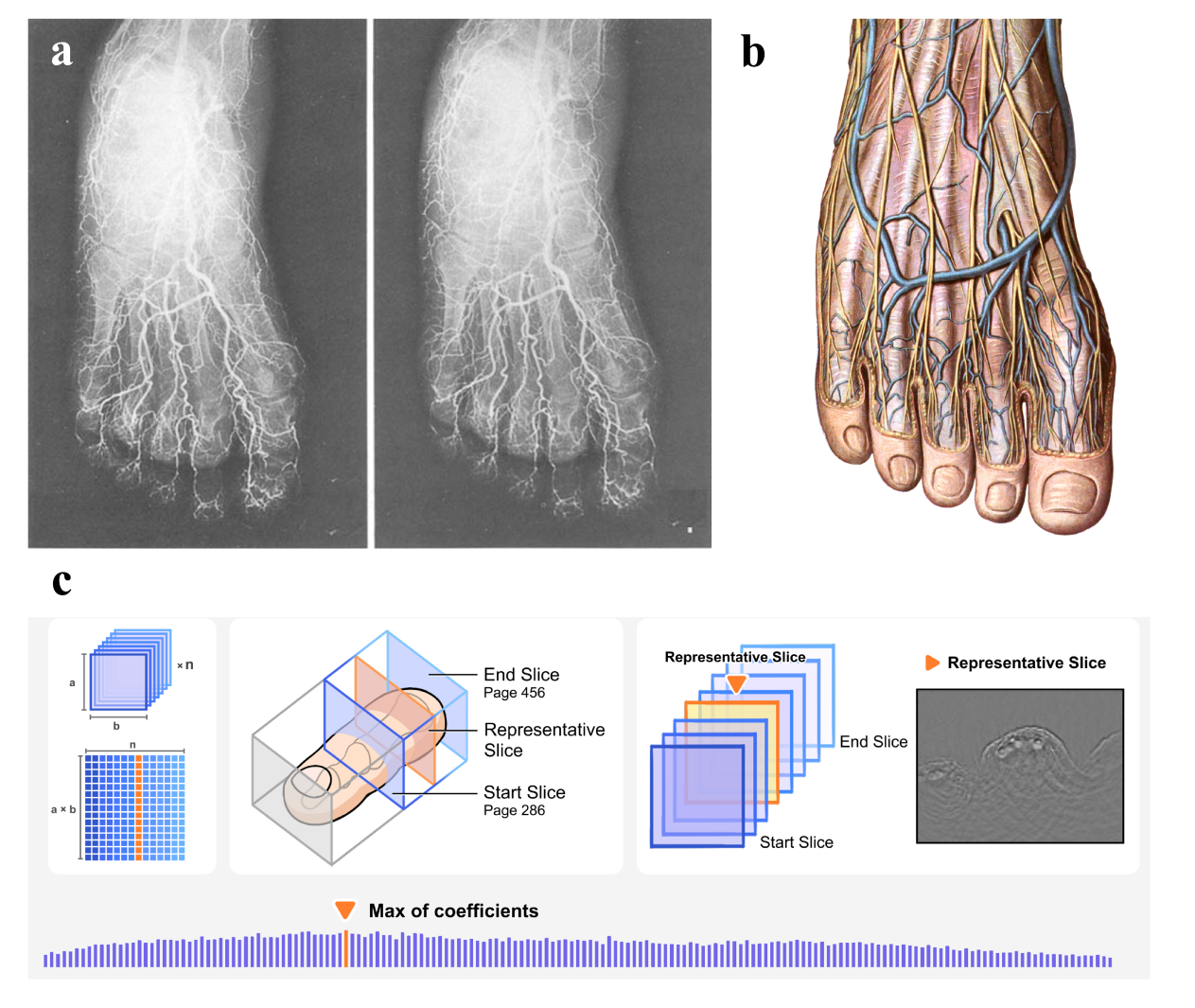
**Supplementary Fig. 7 | Anatomical basis and PCA-assisted selection of the representative second-toe cross-section. a,** Representative vascular imaging of the foot showing the dense and interconnected vascular network extending into the forefoot and toes, supporting toe-level vascular imaging and phenotyping [3]. **b,** Anatomical illustration of the dorsal venous network of the foot, showing longitudinal digital vessels and interconnections across the forefoot and toes [4]. **c,** PCA-assisted representative cross-section selection. Candidate slices were restricted to a predefined anatomical segment of the second toe, vectorized and assembled into a slice matrix. PCA was then used to identify the dominant structural variation, and the slice with the maximum coefficient along the first principal component was selected for PACT-DMS feature extraction. This label-independent procedure reduced subjective slice-selection bias and provided a standardized input for vascular phenotyping.

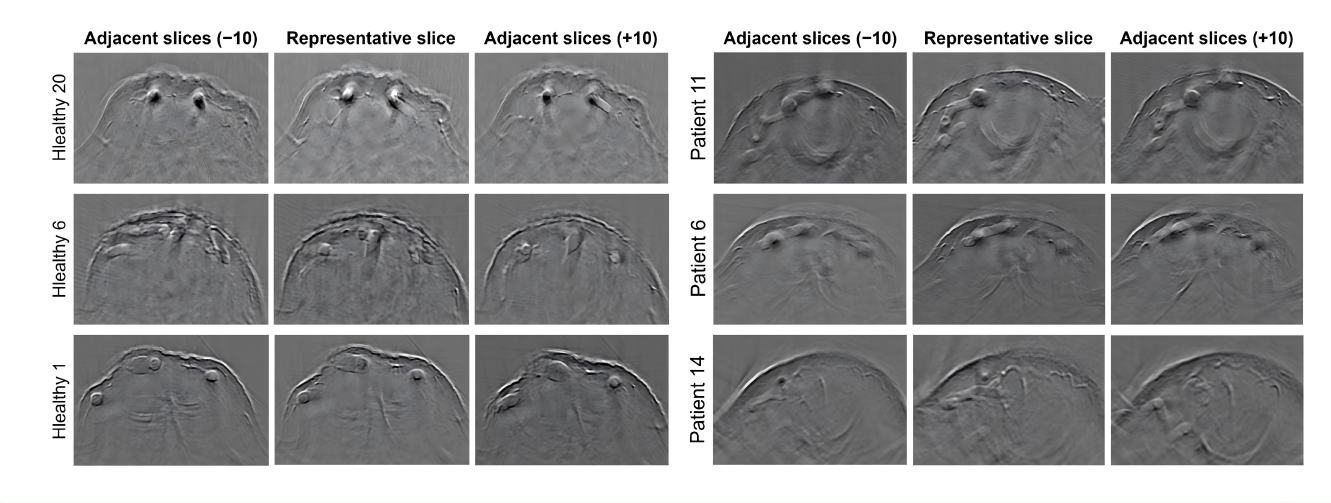

**Supplementary Fig. 8 |** Adjacent-slice consistency analysis of the representative cross-section. To evaluate the sensitivity of representative-section selection to local longitudinal heterogeneity, PACT images of each participant’s representative cross-section and adjacent positions were compared. In each group, three images correspond to the representative section and its adjacent slices, labelled −10, 0 and +10. Under the standardized localization framework used in this study, adjacent slices showed high consistency in overall vascular morphology, trunk-vessel course and local small-vessel distribution, without abrupt structural changes. These findings suggest that the discriminative information of PACT-DMS arises primarily from relatively stable local vascular structural patterns rather than subjective selection of a single “optimal” slice.

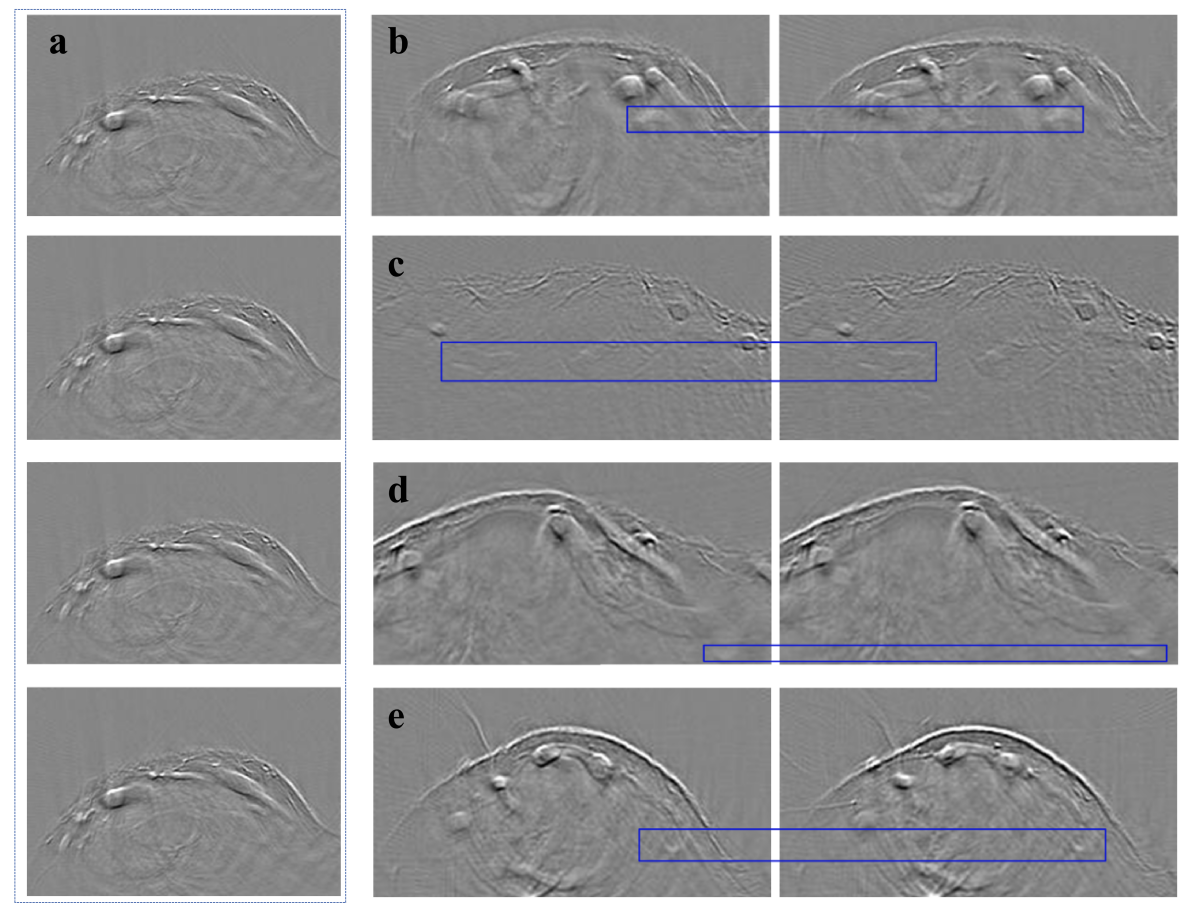

**Supplementary Fig. 9 |** Vessel pulsation scoring criteria and representative examples. a, Example of serial raw PACT images used to illustrate the interpretation of vascular dynamic features between adjacent frames. b, Typical effective pulsation, characterized by global vessel displacement accompanied by changes in calibre or filling state, forming a cardiac-cycle-related expansion–recoil pattern. c, Local shape change, in which the overall vessel position remains stable and only local contour changes are observed; this was not considered definite effective pulsation. d, Intensity variation, in which vessel contour and spatial position are largely unchanged but signal intensity fluctuates, suggesting that the change is mainly brightness-related rather than due to structural periodic displacement. e, No obvious dynamic change, with no definite structural or intensity periodicity other than slight scan-related displacement. In this study, clear global pulsation was considered a positive vessel pulsation feature and was used to construct a candidate perfusion-related dynamic phenotype, rather than as a direct measurement of blood-flow velocity or volume flow.

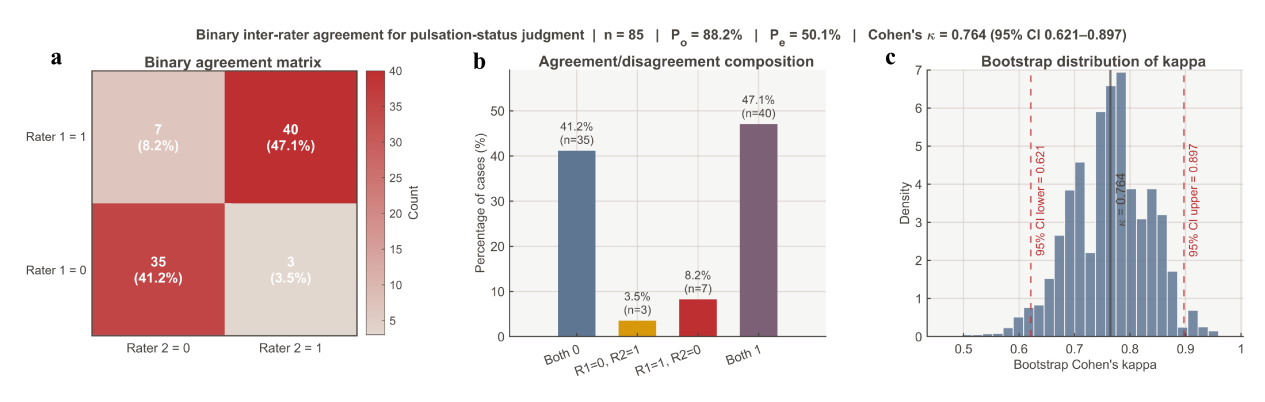

**Supplementar**y Fig. 10 | Interobserver agreement for binary pulsation assessment. a, A 2 × 2 agreement matrix for binary assessment of vessel pulsation by observer 1 and observer 2. Each cell shows the sample count and its proportion of the total sample. **b,** Composition of concordant and discordant assessments, including cases rated negative by both observers, positive by both observers and the two directions of disagreement. **c,** Cohen’s kappa distribution obtained from 5,000 bootstrap resamples. The solid line indicates the point estimate in the original sample, and dashed lines indicate the 95% confidence interval. Among all 85 samples, the overall observed agreement was 88.2%, and Cohen’s kappa was 0.764 (95% CI, 0.628–0.901). These results support good interobserver agreement for binary pulsation assessment under the current rule-based interpretation workflow and provide reproducibility support for using Vessel Pulsation Strength as a candidate dynamic vascular-behaviour feature in the main manuscript.

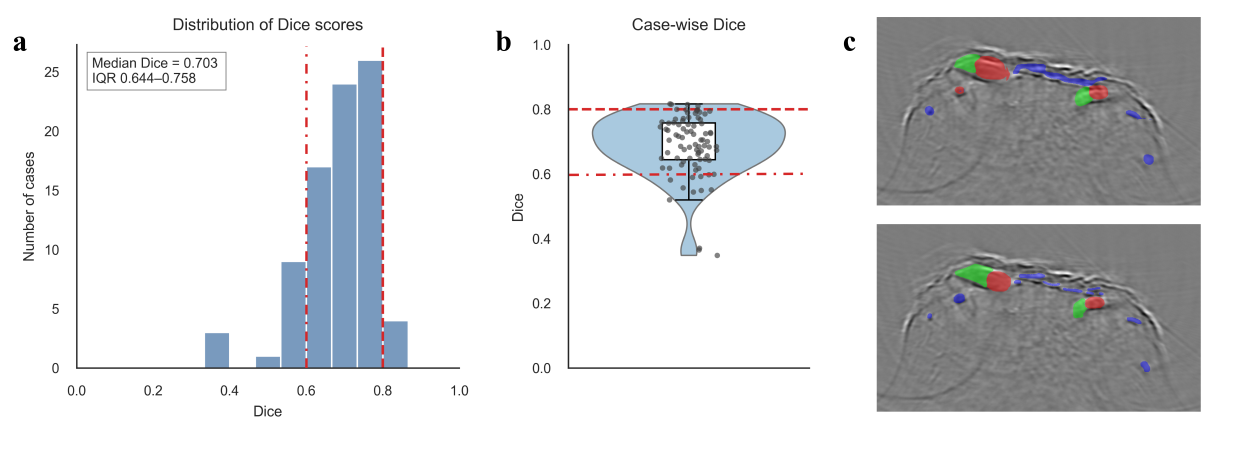
**Supplementary Fig. 11 | Interobserver vascular segmentation consistency analysis and representative example. a,** Histogram of interobserver Dice coefficients across all cases, showing the overall distribution of vascular segmentation consistency and indicating the median, interquartile range and proportion of cases reaching the prespecified reference level. Across all cases, the median Dice coefficient was 0.70 (IQR, 0.64–0.75), and 51.9% of cases had Dice ≥ 0.60. **b,** Case-wise Dice coefficient distribution, showing the range of individual-level agreement and the overall central tendency. **c,** Representative example of two-observer segmentation results (Dice = 0.60). This example illustrates that, in distal toe vascular segmentation, even when two annotations are broadly consistent in trunk-vessel localization, local boundary shifts, inclusion or exclusion of small branches and slight differences in connected vascular regions can reduce the Dice coefficient. Because the target vascular structures are small and have discrete boundaries, a small difference in foreground pixels can have a relatively large effect on Dice, which differs from the interpretation of Dice in large-organ or large-lesion segmentation tasks. The Dice coefficients reported here were calculated using only vascular foreground regions, without including the background class, to avoid overestimation of segmentation agreement caused by the naturally high agreement of large background areas. Together with feature-robustness analysis based on independent annotations by two investigators, these results support acceptable interobserver stability of the current semi-automated vascular annotation workflow and provide methodological support for reproducible PACT-DMS feature extraction.

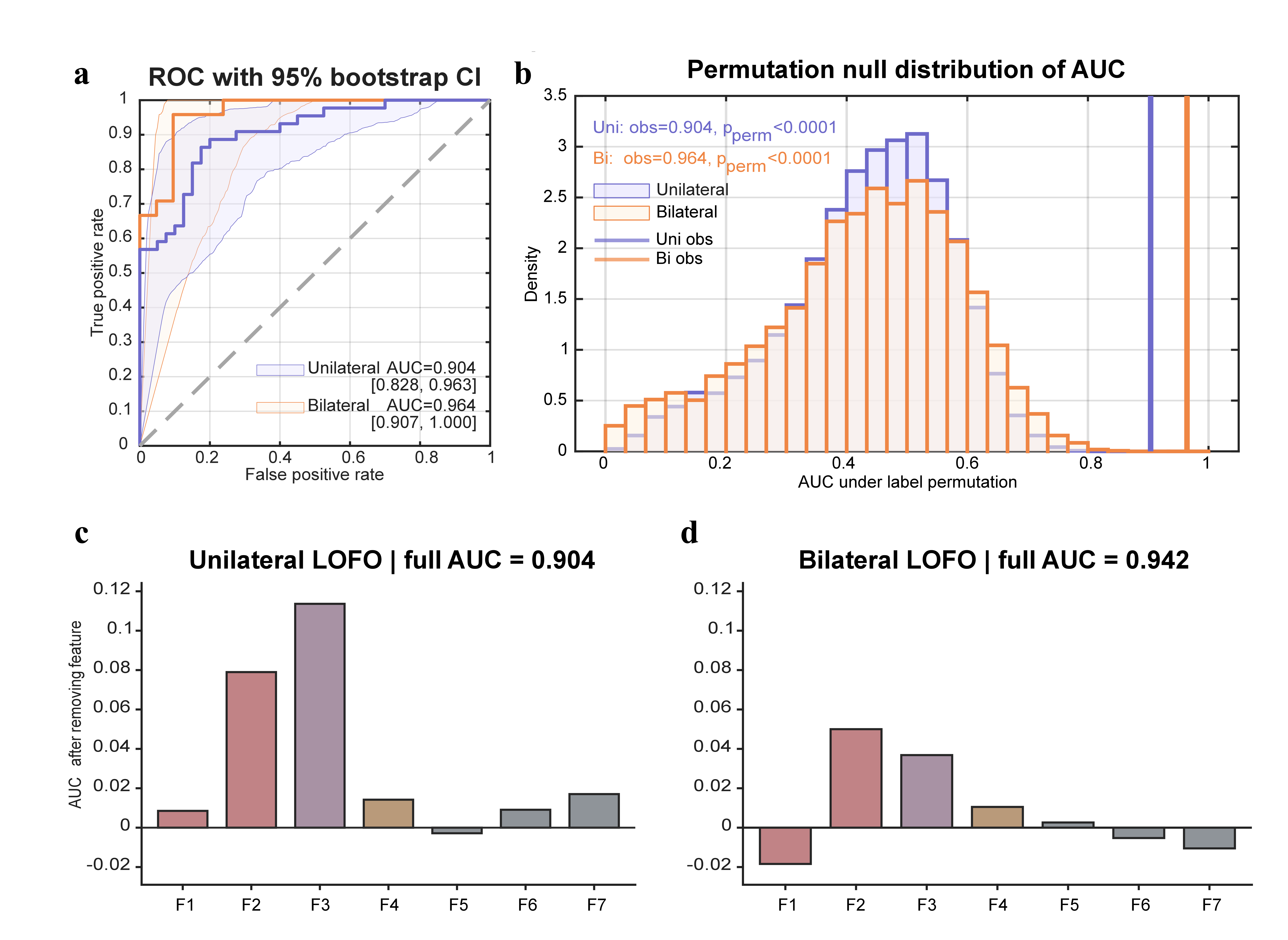

**Supplementary Fig. 12 |** **PACT-DMS model performance, permutation testing and feature ablation analysis. a,** Receiver operating characteristic curves for the unilateral and bilateral models, with 95% confidence intervals estimated by bootstrap resampling. The bilateral fusion model showed higher discriminative performance, with an AUC of 0.964, compared with 0.904 for the unilateral model. **b,** Null distribution of AUC values from label permutation testing. Histograms show the AUC distributions of the unilateral and bilateral models after random label shuffling, and vertical lines indicate the observed AUC under the true labels. The observed AUC values of both models were clearly separated from the main body of the null distributions (permutation test, p < 0.0001), suggesting that the classification performance was unlikely to have arisen from random label assignment. **c,d,** Leave-one-feature-out (LOFO) ablation analysis for the unilateral and bilateral models, respectively. The y axis shows the change in model AUC after removing each feature; larger values indicate a stronger unique contribution of that feature to discriminative ability, whereas values close to zero or below zero suggest limited contribution or partial redundancy with other features. F2 and F3 showed notable marginal contributions in both models, whereas F5–F7 showed relatively smaller marginal contributions. In the bilateral model, removal of some features slightly increased AUC, suggesting that feature redundancy may increase after bilateral fusion. This analysis supports the interpretation that the discriminative ability of PACT-DMS does not rely entirely on a single feature but arises from the combination of multiple interpretable vascular phenotypes, while also indicating that feature contributions differ between unilateral and bilateral models.

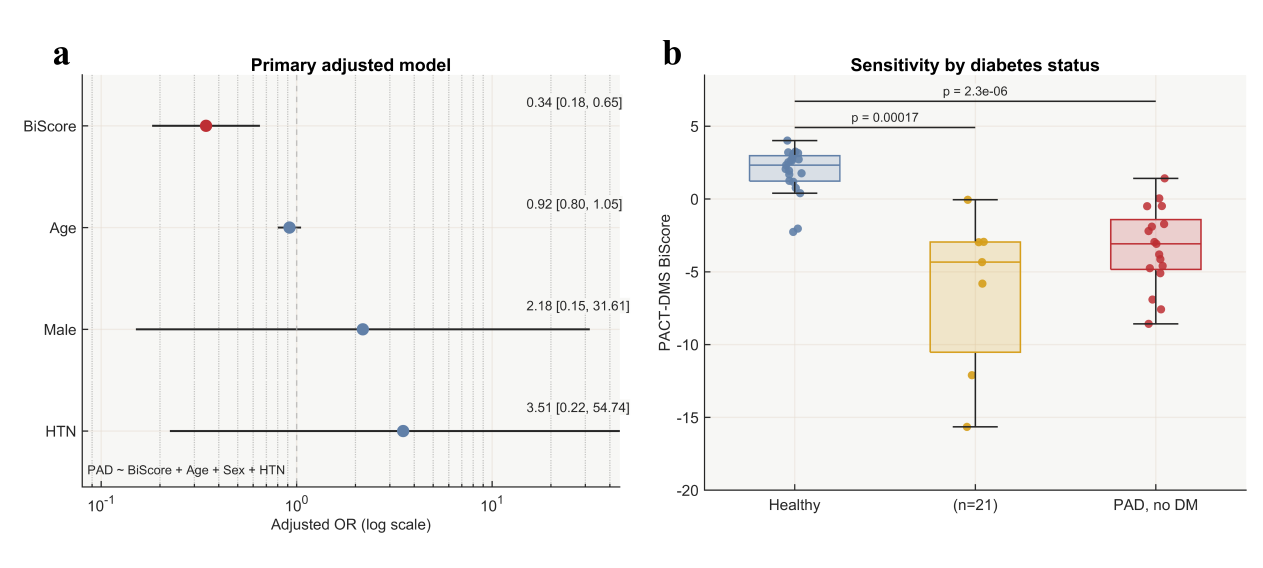

**Supplementary Fig. 13 | Cohort covariate and diabetes-stratified sensitivity analyses. a,** Subject-level logistic regression analysis with PAD status as the dependent variable, BiScore as the primary imaging variable and adjustment for age, sex and hypertension. After limited covariate adjustment, BiScore remained associated with PAD status, suggesting that the association between PACT-DMS-related phenotypes and PAD was not fully explained by these basic clinical variables. **b,** Sensitivity analysis stratified by diabetes status. Because diabetes is an important upstream vascular risk factor closely related to PAD, diabetes was controlled during healthy-control recruitment to reduce overt metabolic vascular confounding. Both the non-diabetic PAD subgroup and the PAD subgroup with diabetes showed lower BiScore than healthy controls, with changes in a direction consistent with PAD-related distal vascular phenotypic abnormality. This analysis supports the rationale for the cohort design and suggests that the imaging differences observed in the main manuscript are not solely dependent on differences in diabetes distribution. Owing to the limited sample size, this covariate analysis should be interpreted as a sensitivity analysis rather than a fully adjusted confounding-control model.

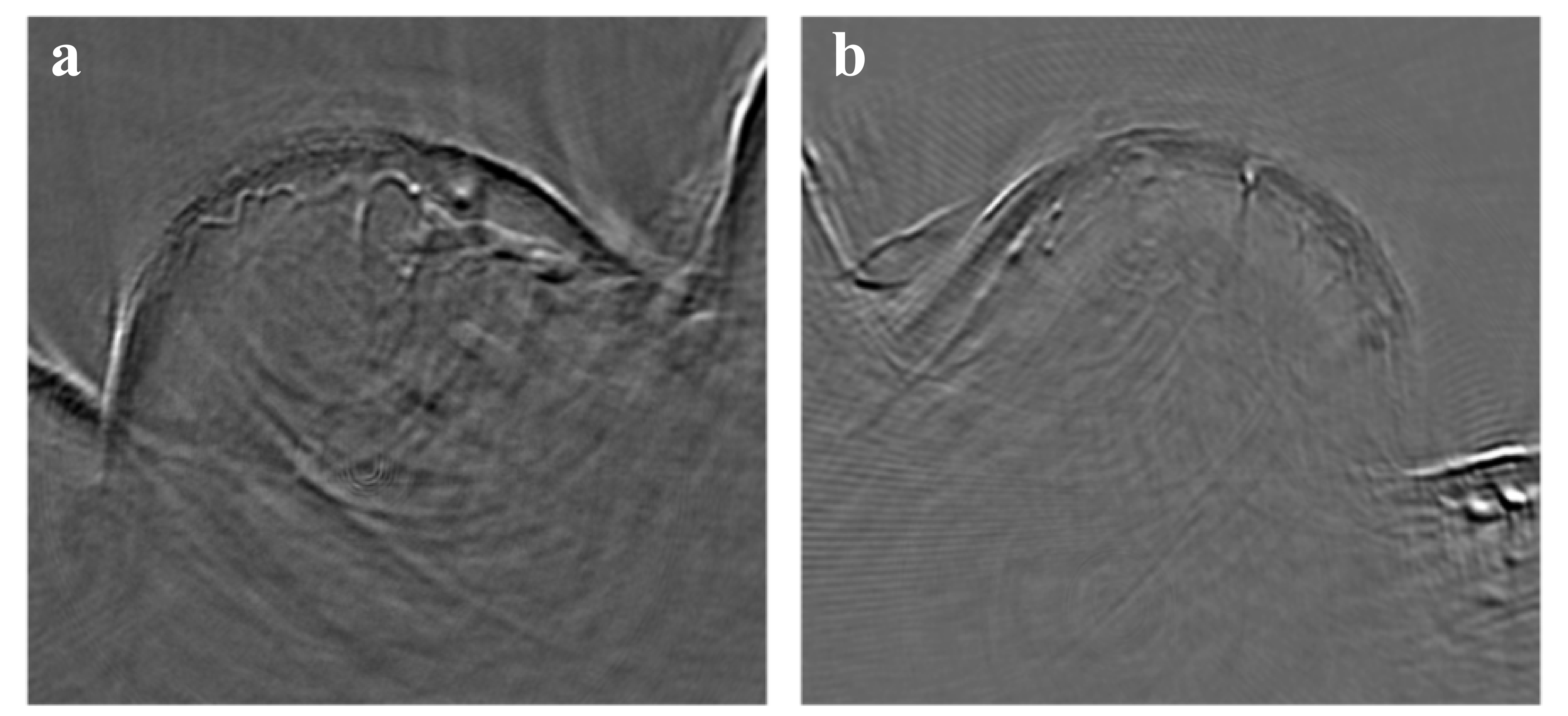

**Supplementary Fig. 14 | Example of bilateral distal toe vascular phenotypes in Patient 14. a,** Representative PACT cross-sectional image of the left foot of Patient 14. The left foot shows marked sparsity of distal vascular structures, insufficient visualization of trunk vessels and residual, focal small-vessel structures. The PACT-DMS score of this foot was used for the preoperative distal vascular phenotype in the main manuscript. **b,** Representative PACT cross-sectional image of the right foot of Patient 14. The right foot shows a more visually severe impairment of the distal vascular bed, characterized mainly by near absence of longitudinal trunk vessels and abnormal residual small-vessel structures. Because the right foot showed an extreme phenotype with almost complete absence of L-type trunk vessels, assigning an even lower score directly on the basis of this extreme morphology could introduce an unstable estimate driven by a single case and could overstate the scoring effect in the case illustration. Therefore, in the main and supplementary analyses, the right-foot image is retained as qualitative evidence of severe distal vascular phenotype, whereas the labelled score follows the PACT-DMS score of the same patient’s left foot used in the main quantitative analysis, as a conservative patient-level phenotypic representation. Overall, the bilateral images of Patient 14 indicate severe structural impairment of the distal vascular bed. The left foot was used for quantitative scoring and case comparison in the main manuscript, whereas the right foot serves as key imaging evidence supporting extensive bilateral distal microcirculatory abnormality in this patient. The right foot was not included as an additional independent limb-level sample in model training or performance evaluation, but was used for postoperative recovery assessment.

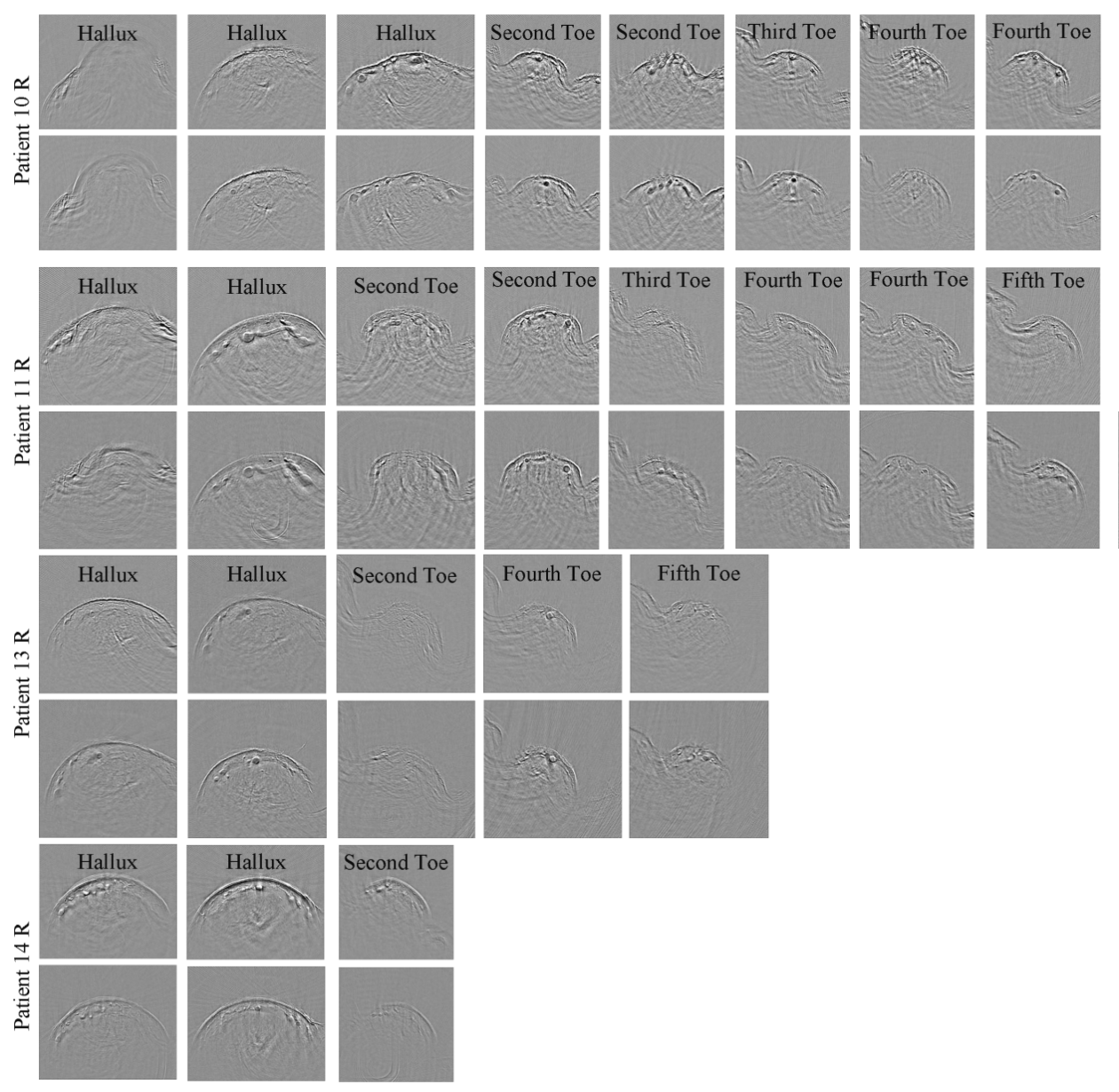

**Supplementary Fig. 15 | Post-revascularization changes in distal vascular phenotype in Patients 10–14.** Multi-toe PACT cross-sectional images before and after revascularization are shown for Patients 10–14 to assess postoperative changes in distal vascular filling, small-vessel visibility and local vascular structural display. Postoperative change was described according to the proportion of acquired toes on the operated side showing visible vascular phenotypic changes: ≥60% was defined as marked change, 20–60% as moderate change and <20% as limited change. This figure supports the interpretation in the main manuscript that postoperative PACT phenotypes may provide exploratory imaging clues about distal vascular-state changes after revascularization, rather than establishing a standardized treatment-response scoring model.

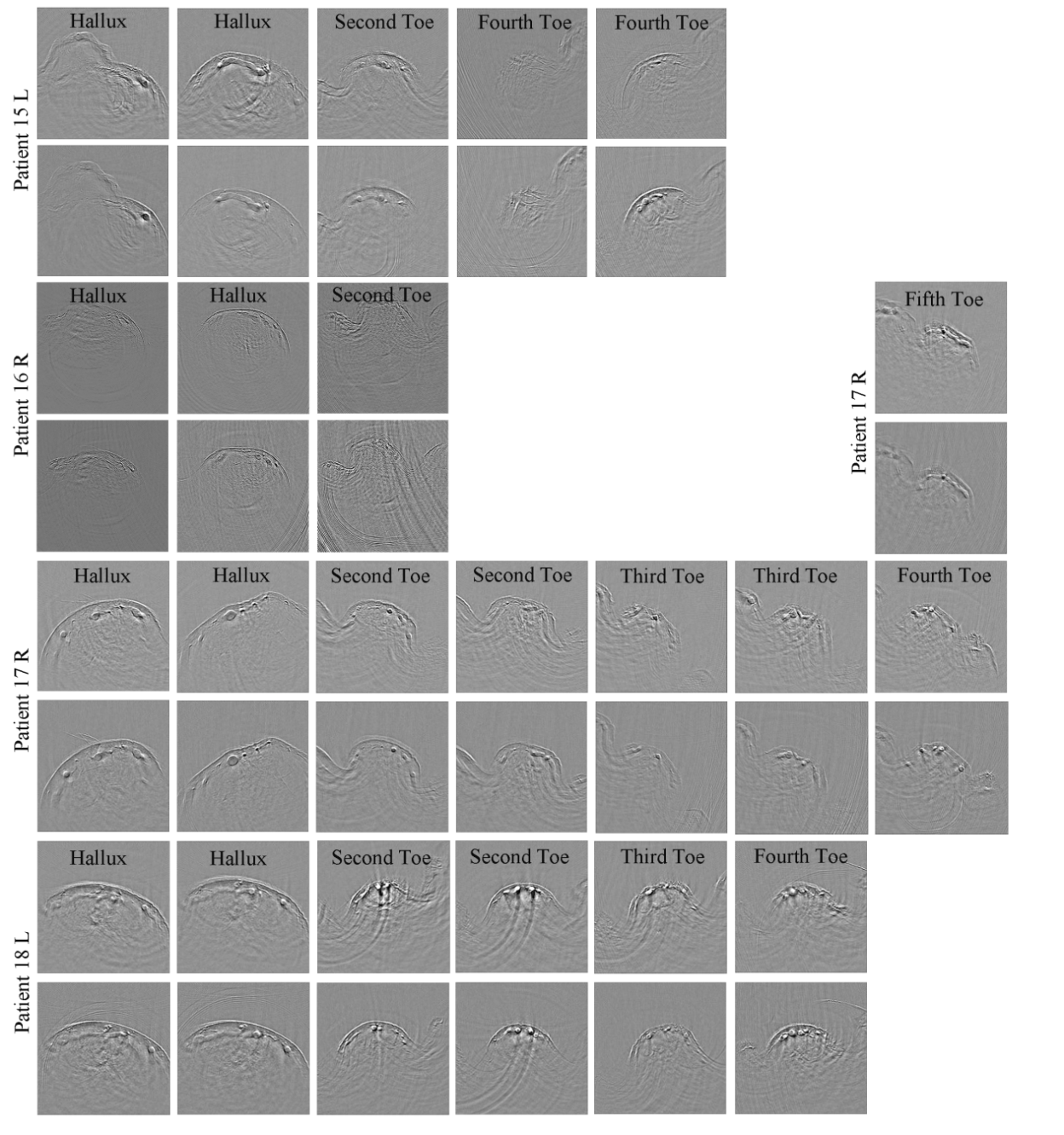

**Supplementary Fig. 16 |** **Post-revascularization changes in distal vascular phenotype in Patients 15–18.** Multi-toe PACT cross-sectional images before and after revascularization are shown for Patients 15–18. Cases are described according to the proportion of toes showing visible vascular phenotypic changes, illustrating interindividual variation in postoperative distal vascular filling and small-vessel visibility. This figure complements the representative cases in Fig. 6 of the main manuscript and further indicates that pressure-based ABI improvement and recovery of distal vascular phenotype may not be fully synchronous, as the two reflect different dimensions of vascular status.

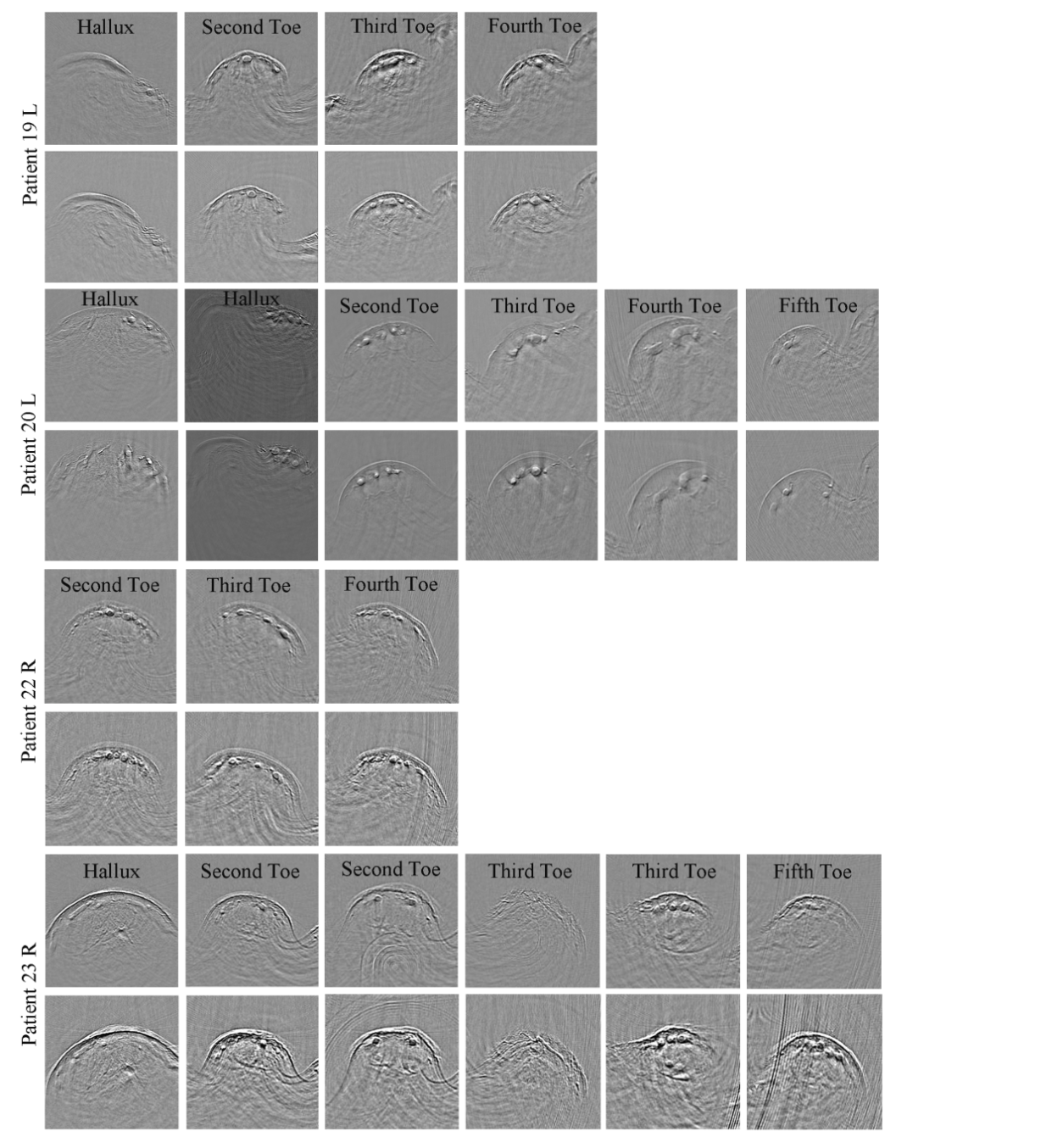

**Supplementary Fig. 17 |** **Post-revascularization changes in distal vascular phenotype in Patients 19–23.** Multi-toe PACT cross-sectional images before and after revascularization are shown for Patients 19–23, illustrating postoperative changes in distal vascular filling, small-vessel visibility, and vascular structure. Together with Supplementary Figs. 15 and 16, these cases provide individual-level evidence supporting the exploratory postoperative analysis, demonstrating that PACT can visualize dynamic changes in peripheral microcirculation after revascularization. These analyses are intended as qualitative and semiquantitative assessments rather than a standardized postoperative treatment-response model.

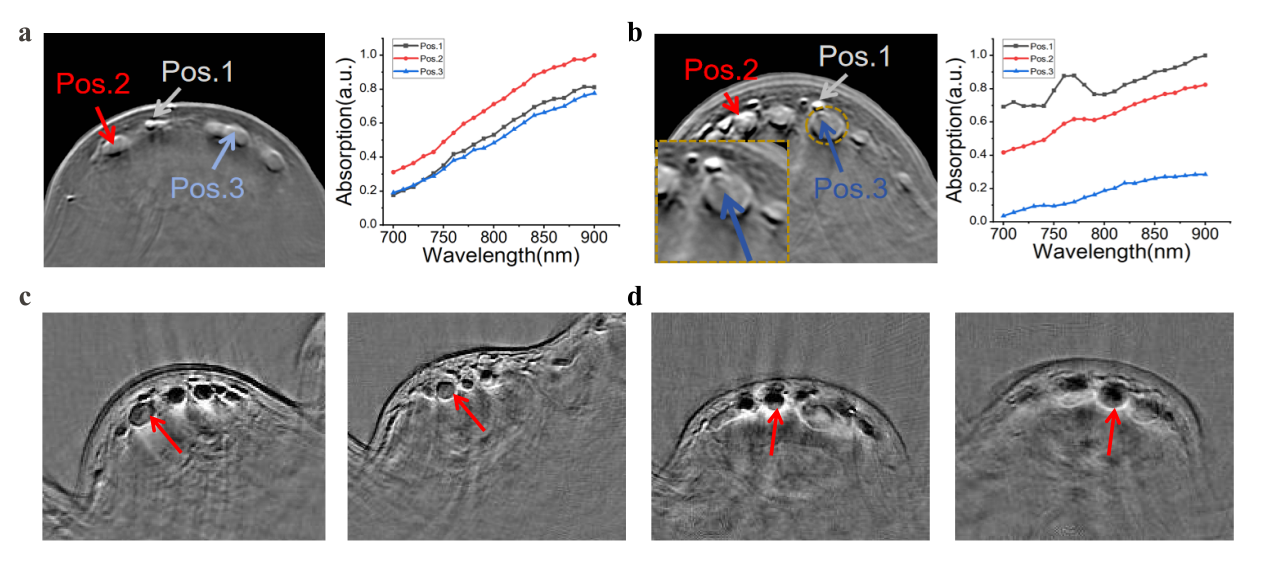

**Supplementary Fig. 18 | Exploratory local vascular signal heterogeneity revealed by multispectral PACT. a,** Representative multispectral photoacoustic signals and corresponding spectral profiles of small vessels in a healthy volunteer. The spectral pattern of these small vessels generally resembles that of oxygenated hemoglobin. **B,** Representative multispectral photoacoustic signals and corresponding spectral profiles of small vessels in a patient with PAD. Some small-vessel signals exhibit spectral features more consistent with deoxygenated hemoglobin. In addition, reduced signal intensity is observed in some larger vessels, which may reflect decreased intravascular hemoglobin content, increased contribution from non-hemoglobin components, or alterations in the composition of intravascular contents. **c,d,** Representative images from two patients with poor postoperative recovery. Heterogeneous, flocculent intravascular signal patterns can be observed within some vessels, which are more clearly appreciated under the inverted color scale. These exploratory observations suggest that multispectral PACT, together with morphological assessment of intravascular signal patterns, may provide clues for further investigation of oxygenation status, vascular contents, and the local tissue microenvironment in the distal vascular bed of PAD. However, these findings are currently based on only a small number of cases and lack independent validation by biochemical, pathological, or hemodynamic reference standards. Therefore, they should be regarded as exploratory and hypothesis-generating, rather than interpreted as validated markers of blood oxygenation, biochemical composition, or prognosis.

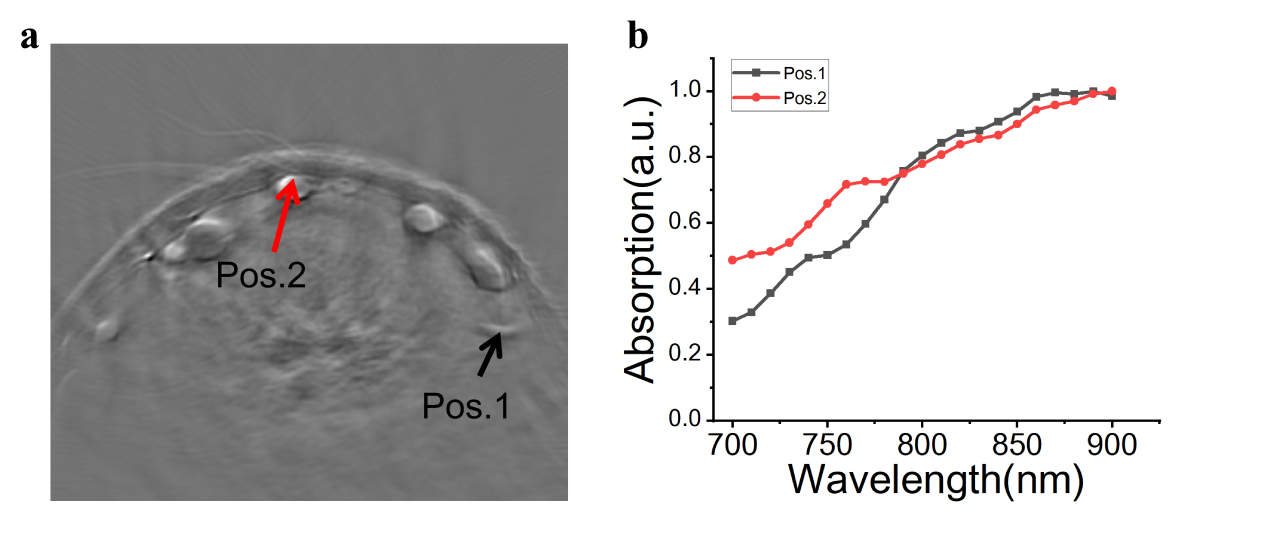

**Supplementary Fig. 19 |** **Challenges in spectrally distinguishing arteries and veins in distal toe vessels. a,** Representative multispectral PACT cross-sectional image of a distal toe. Pos. 1 indicates a vessel with morphology more consistent with an arterial structure, whereas Pos. 2 indicates a superficial vessel morphologically consistent with a vein. **b,** Normalized multispectral photoacoustic absorption profiles extracted from Pos. 1 and Pos. 2. Although PACT can in principle use oxygenation-related spectral differences to help distinguish arteries from veins, the two positions showed partially overlapping spectral trends under the imaging conditions used in this study. These observations indicate that stable arteriovenous classification at the single-vessel level based solely on multi-wavelength spectral behaviour remains challenging in the distal toe vascular bed. Local optical attenuation, vessel depth, superficial tissue staining and mixed vascular contents may all affect spectral interpretation. Therefore, the PACT-DMS analysis in the main manuscript assessed the distal toe vascular bed as an integrated structural–dynamic phenotype and did not require stable arteriovenous classification of each individual vessel.

### Supplementary Tables

| **Characteristic** | **PAD group (n = 24)** | **Healthy control group (n = 21)** | **P value** |
| --- | --- | --- | --- |
| Age, years | 68.2 ± 9.1 | 68.1 ± 5.3 | 0.961 |
| Male sex, n (%) | 14 (58.3%) | 11 (52.4%) | 0.769 |
| Diabetes mellitus, n (%) | 17 (70.8%) | 0 (0.0%) | <0.001 |
| Hypertension, n (%) | 19 (79.2%) | 9 (42.9%) | 0.016 |
| Hyperlipidaemia, n (%) | 7 (29.2%) | 6 (28.6%) | 1.000 |
| Coronary artery disease, n (%) | 9 (37.5%) | 3 (14.3%) | 0.101 |
| History of myocardial infarction or stroke, n (%) | 6 (25.0%) | 2 (9.5%) | 0.252 |
| Renal insufficiency, n (%) | 6 (25.0%) | 0 (0.0%) | 0.023 |
| Minimum ABI, median [IQR] | 0.47 [0.30–0.66] | NA | — |
| Data are presented as mean ± SD, n (%), or median [IQR], as appropriate. | | | |
| P values were calculated using Welch’s t test for age and Fisher’s exact test for categorical variables. | | | |
| ABI, ankle-brachial index; IQR, interquartile range; PAD, peripheral artery disease. | | | |
| ABI was not assessed in healthy controls. | | | |

**Supplementary Table 1 |** Baseline characteristics of the study cohort. The table summarizes demographic and clinical characteristics of patients with PAD and age-matched healthy controls, including age, sex, diabetes, hypertension, hyperlipidaemia, coronary artery disease, history of myocardial infarction or stroke and renal insufficiency. This table describes the cohort composition and provides clinical context for the PAD-versus-healthy-control analyses in the main manuscript.

| **Case No.** | **Clinical presentation at admission** | **1-month telephone follow-up** | **6-month telephone follow-up** |
| --- | --- | --- | --- |
| P1 | Chronic bilateral intermittent claudication with recent worsening of right-sided pain and numbness, accompanied by rest pain. | At follow-up, pain and numbness had resolved and nocturnal rest pain was absent; intermittent lower-extremity edema persisted. | At follow-up, intermittent coldness and occasional rest pain persisted; further vascular management was under consideration. |
| P6 | Exertional lower-extremity swelling and pain, more pronounced on the right, with recent intermittent claudication. | At follow-up, walking capacity had markedly improved, swelling had resolved, and occasional numbness remained. | At follow-up after additional vascular procedures, walking-related swelling, pain, and bilateral groin numbness persisted. |
| P13 | Long-standing bilateral lower-extremity swelling and pain with recent worsening. | At follow-up, pain and numbness had resolved; occasional edema remained. | At follow-up, erythema, swelling, and ulceration had resolved; less frequent and less severe nocturnal rest pain remained, with improved sleep. |
| P14 | Right forefoot pain, redness, swelling, and interdigital ulceration. | At follow-up, pain was reduced and limited to occasional nocturnal episodes. | At follow-up, pain and swelling had resolved, with good recovery. |
| P15 | Long-standing intermittent claudication of the left lower extremity with soreness and pain. | At follow-up, pain, soreness, and numbness had resolved, with improved walking capacity. | At follow-up, left-foot ulceration had resolved and claudication had improved, with substantially increased walking distance. |
| P18 | Long-standing left lower-extremity pain and foot numbness with recent worsening. | At follow-up, pain and numbness had resolved, with occasional soreness after prolonged walking. | At follow-up, pain was relieved; exertional foot fullness and mild nocturnal numbness remained. |

**Supplementary Table 2 |** Clinical presentation and follow-up information of patients with PAD. The table lists the major clinical presentations, postoperative follow-up information and symptom changes of patients with PAD, supporting the exploratory perioperative analysis in the main manuscript. It provides individual-level clinical context for interpreting potential relationships among preoperative PACT-DMS phenotype, postoperative Rutherford-class changes and postoperative PACT imaging phenotypes.

| **Case No.** | **Operated limb** | **Preoperative ABI, left** | **Preoperative ABI, right** | **Postoperative ABI, left** | **Postoperative ABI, right** |
| --- | --- | --- | --- | --- | --- |
| P10 | Right | 0.51 | 0.62 | 0.88 | 0.75 |
| P11 | Right | <0.3 | <0.3 | <0.3 | 0.79 |
| P13 | Right | 1.02 | 0.67 | 0.97 | 1.01 |
| P14 | Right | 0.52 | <0.3 | 0.52 | 0.94 |
| P15 | Left | 0.67 | <0.3 | 0.9 | 0.56 |
| P16 | Right | 0.66 | 0.83 | 0.83 | 0.88 |
| P17 | Right | 1.14 | <0.3 | 1.14 | 1.11 |
| P18 | Left | 0.54 | 0.72 | 0.95 | 0.92 |
| P19 | Left | <0.3 | 0.76 | 0.9 | 0.85 |
| P20 | Left | 0.78 | 1.16 | 0.89 | 1.22 |
| P22 | Right | 0.63 | 0.38 | 0.44 | 0.67 |
| P23 | Right | 0.74 | 0.74 | 0.81 | 0.66 |
| Abbreviation: ABI, ankle-brachial index. Values were translated directly from the source worksheet and reformatted for journal submission; no statistical analyses were performed. | | | | | |

**Supplementary Table 3 |** Preoperative and postoperative ABI values in patients undergoing revascularization. The table lists the operated limb and preoperative and postoperative ABI values for the left and right feet in patients who underwent revascularization and completed ABI testing before and after surgery. It supports the exploratory postoperative analysis in the main manuscript, particularly cases in which ABI changed only modestly but PACT images showed visible distal vascular phenotypic changes. ABI, ankle-brachial index.

| **Method** | **True positives** | **False negatives** | **Total PAD limbs** | **Sensitivity (%)** |
| --- | --- | --- | --- | --- |
| ABI | 35 | 9 | 44 | 79.5% |
| PACT-DMS | 37 | 7 | 44 | 84.1% |
| Abbreviations: ABI, ankle-brachial index; PACT-DMS, photoacoustic computed tomography-derived microcirculatory score. | | | | |
| TP indicates true positives; FN, false negatives. Sensitivity was calculated as TP / (TP + FN). | | | | |
| In this exploratory limb-level comparison, abnormal ABI was defined as ABI < 0.90 and non-abnormal ABI as ABI ≥ 0.90. | | | | |

**Supplementary Table 4 |** Classification results of ABI and PACT-DMS for PAD limbs. The table compares ABI and PACT-DMS classifications in PAD limbs with available ABI records, including true positives, false negatives, total number of PAD limbs and sensitivity. This table supports the ABI-insensitive subgroup analysis in the main manuscript by showing that PACT-DMS and ABI classifications of PAD limbs are not completely concordant. The result is intended only to explore whether distal photoacoustic microcirculatory phenotypes provide complementary information beyond pressure-based ABI and should not be interpreted as definitive validation of PACT-DMS superiority over ABI.

| **Feature domain** | **Feature code** | **Feature name** | **Definition** | **Pathophysiological interpretation** |
| --- | --- | --- | --- | --- |
| Morphological feature | F1 | Maximum L-vessel area | Reflects the calibre of the most prominent longitudinal trunk in the cross-section, defined as the cross-sectional area of the largest L-type vessel. | A larger area theoretically indicates greater blood-carrying capacity per unit time and provides an intuitive indicator for assessing whether the local trunk is markedly narrowed or collapsed. |
| Morphological feature | F2 | Maximum L-vessel circularity | Reflects the morphological integrity of the trunk cross-section, defined as the logarithmic transformation of classical circularity, F_2_=log(4πA/P^2^), calculated from the largest L-type vessel component. | Under normal conditions, distal arterial cross-sections are expected to be relatively circular, yielding F2 values closer to 0. Flattened, elongated or irregular cross-sections yield more negative F2 values, suggesting impaired trunk-vessel morphological integrity. |
| Functional feature | F3 | Vessel pulsation strength | Represents a rule-based binary pulsation phenotype derived from time-series PACT data, indicating whether definite global pulse-like cyclic vessel motion is present. | Clear and large-amplitude pulsation suggests that the vessel segment remains well coupled to the central arterial system and has relatively preserved perfusion function. Markedly weakened or absent pulsation suggests restricted local blood flow or reduced vascular compliance. |
| Compensatory feature | F4 | Maximum S-vessel cluster area | Identifies the largest S-type small-vessel cluster in the cross-section by spatial clustering and calculates its total area. | Can be interpreted as an estimate of the scale of compensatory transport pathways that emerge when the trunk is limited. A larger cluster suggests more evident focal microcirculatory remodelling and may indirectly reflect the degree of upstream supply limitation. |
| Distributional feature | F5 | T-L vessel count difference | Measures collateral expansion relative to the trunk as the number of transverse (T-type) vessels minus the number of longitudinal (L-type) vessels. | In healthy individuals, this difference is usually close to 0, indicating a stable trunk-to-collateral ratio. In PAD, the value may increase owing to collateral proliferation or decrease owing to global destruction, reflecting heterogeneous remodelling patterns across patients. |
| Distributional feature | F6 | S-vessel count | Counts all small vessels meeting the threshold in the entire image, representing the global level of microcirculatory proliferation or loss. | Unlike the maximum cluster captured by F4, F6 emphasizes the overall small-vessel burden across different regions and serves as a global complement to the local clustering signal. |
| Distributional feature | F7 | Trunk-small vessel junction count | Counts visible junctions between L/T-type vessels and S-type small vessels within the cross-section. | Reflects the topological coupling between trunk vessels and the microcirculatory network. More junctions suggest a relatively intact trunk-branch-capillary continuum; fewer junctions indicate decoupling between trunks and the microcirculation, which may be unfavourable for blood-flow redistribution and compensation. |
| Abbreviations: PACT, photoacoustic computed tomography; PAD, peripheral artery disease. | | | | |
| Definition notes: L-type vessels indicate longitudinal trunk vessels; T-type vessels indicate transverse/collateral vessels; S-type vessels indicate small vessels. | | | | |
| Junction refers to a visible connection point between a trunk vessel and a small vessel within the cross-section. | | | | |

**Supplementary Table 5 |** Definitions and potential physiopathological implications of seven PACT-based vascular imaging features. The table lists the seven semantic vascular imaging features used in PACT-DMS, including maximum L-vessel area, maximum L-vessel circularity, vessel pulsation strength, maximum S-vessel cluster area, T-L vessel count difference, S-vessel count and trunk-small vessel junction count. Each feature corresponds to a specific dimension of vascular morphology, microcirculatory remodelling, topological coupling or dynamic vascular behaviour, supporting the methodological positioning of PACT-DMS as an interpretable distal microcirculatory phenotype score in the main manuscript. PACT, photoacoustic computed tomography; PAD, peripheral artery disease; L-type vessels indicate longitudinal trunk vessels; T-type vessels indicate transverse or collateral vessels; S-type vessels indicate small-vessel plexuses; junction refers to a visible connection point between a trunk vessel and a small vessel within the cross-section.

| **Item** | **Setting** | **Rationale or leakage-control note** |
| --- | --- | --- |
| Classifier and inputs | Linear support vector machine using the seven predefined PACT-DMS features; linear kernel; no supervised feature selection. | Low-capacity model chosen to preserve interpretability and feature traceability. |
| Regularization and hyperparameters | Box constraint **C = 1**, fixed for all cross-validation folds; hyperparameter tuning = **none**. | No model hyperparameter was estimated using the held-out participant. |
| Class weighting | **None**. | No class-weighting strategy was estimated from the full dataset or from the held-out participant. |
| Feature standardization | z-score standardization; mean and s.d. estimated from training participants in each fold and then applied to the held-out participant. | Prevents information leakage from the held-out participant during preprocessing. |
| Cross-validation unit | Subject-level leave-one-out cross-validation; all available limbs from the held-out participant were excluded from training. | Prevents left-right bilateral samples from the same participant appearing in both training and testing. |
| Decision score | Continuous SVM decision function for the healthy-reference class; lower scores indicate a more PAD-like phenotype and higher scores indicate a healthy-reference phenotype. | Score orientation was fixed for interpretation and used consistently across unilateral and bilateral analyses. |
| Binary threshold | Default zero threshold of the SVM decision function. | No supervised decision threshold was estimated using the held-out participant. |
| Bilateral fusion | Left- and right-foot decision scores from the held-out participant were averaged before thresholding when both sides were available. | Fusion was applied only after held-out limb-level scores had been generated. |
| Internal robustness analyses | Bootstrap uncertainty estimation with **1,000** resamples; participant-level permutation testing with **10,000** iterations; leave-one-feature-out ablation; Spearman feature-correlation analysis with FDR correction. | Used to assess uncertainty, chance-level separation, feature dependence, feature redundancy and robustness to key participant-level covariates. |
| Implementation | MATLAB R2025a; linear SVM implemented using fitcsvm; ROC/AUC estimated using perfcurve; custom MATLAB scripts were used for subject-level LOOCV, bootstrap resampling, permutation testing and feature ablation. | Software details and analysis code availability should be reported for reproducibility. |
| Clinical positive class for performance reporting | PAD | Although the continuous SVM decision score was oriented toward the healthy-reference phenotype, sensitivity, specificity, ROC/AUC and related clinical performance metrics were reported with PAD treated as the positive class. |

**Supplementary Table 6** | Classifier implementation details for PACT-DMS. The table summarizes the implementation settings of the PACT-DMS diagnostic classifier, including the input features, linear support vector machine configuration, regularization parameter, class-weighting strategy, feature standardization, decision-threshold definition, bilateral fusion rule and cross-validation procedure. It supports the diagnostic modelling analysis in the main manuscript by clarifying how the seven predefined vascular features were converted into continuous PACT-DMS decision scores and binary classification outputs. All feature standardization, model fitting, parameter setting and thresholding procedures were performed without access to the held-out participant during subject-level leave-one-out cross-validation. This table is not intended to introduce an additional model or post hoc optimization procedure, but to provide reproducibility details and leakage-control information for interpreting the reported PACT-DMS performance.

| **Patient ID** | **Left**  **/right** | **ABI** | **PACT-DMS score** | **Major abnormal features** | **Corresponding imaging/symptom notes** |
| --- | --- | --- | --- | --- | --- |
| P2 | L | 1.17 | −5.550 | F4↓, F2↓, F6↓ | Left-foot ulceration with poor healing. CTA showed bilateral multilevel calcific arterial disease with more pronounced left-sided distal involvement. |
| P2 | R | 1.07 | 0.980 | F6↓, F7↓ | CTA showed bilateral multilevel calcific arterial disease with more pronounced right distal-foot involvement; however, the local PACT score of the selected toe did not indicate marked abnormality. |
| P6 | L | 0.9 | −3.750 | F2↓, F3↓, F7↓ | Intermittent claudication. CTA showed multilevel left lower-limb arterial stenotic disease with impaired bilateral distal runoff. |
| P8 | L | 0.95 | −3.490 | F3↓, F4↓, F7↓ | History of lower-limb revascularization with persistent exertional symptoms. CTA showed postoperative graft-related and bilateral multilevel distal arterial abnormalities, with greater clinical severity on the contralateral side. |
| P12 | L | 1.08 | −1.880 | F3↓, F6↓, F4↓ | Intermittent claudication of the right lower extremity. CTA showed bilateral multilevel distal arterial disease with impaired distal runoff. |
| P13 | L | 1.02 | −1.657 | F2↓, F6↓, F4↓ | Bilateral lower-extremity swelling and pain, more pronounced on the left. CTA showed multilevel aortoiliac and femoral disease with impaired bilateral distal runoff. |
| P17 | L | 1.14 | 1.580 | F2↓, F6↓, F4↓ | Clinically severe disease was greater on the right. CTA showed multilevel right-sided occlusive disease and bilateral distal runoff impairment, with only mild left-sided disease. |
| P20 | R | 1.16 | −2.160 | F3↓, F2↓ | Intermittent claudication with bilateral exertional symptoms. CTA showed bilateral distal arterial disease and impaired pedal runoff. |

**Supplementary Table 7 |** PACT-DMS scores, major abnormal features and clinical imaging notes in ABI-normal or selected key cases. The table lists ABI, PACT-DMS score, major abnormal features and corresponding clinical manifestations or CTA/DSA imaging notes for selected cases and sides. It supports the ABI-insensitive limb analysis in the main manuscript by helping to interpret the clinical and imaging background corresponding to different combinations of abnormal features in PACT-DMS-positive cases. This table is not intended to establish a new diagnostic threshold or prognostic model, but to provide individual-level supplementary material for interpreting heterogeneity in PACT-DMS phenotypes.

### Supplementary Data Descriptions

Supplementary Data 1 | Subject baseline characteristics. This worksheet provides de-identified subject-level baseline information, including randomly assigned study ID, group, non-overlapping 5-year age range, sex, left and right ABI, diabetes, hypertension, hyperlipidaemia, coronary artery disease, history of myocardial infarction or stroke and renal insufficiency. ABI classification rule: For the exploratory limb-level comparison between ABI and PACT-DMS classification, abnormal ABI was defined as ABI < 0.90, and non-abnormal ABI was defined as ABI ≥ 0.90. This rule was used to classify the 44 PAD limbs with available ABI records into abnormal-ABI and non-abnormal-ABI groups. These data provide the source data for the cohort description in the main manuscript, Supplementary Table 1 and the covariate/diabetes-stratified sensitivity analyses.

Supplementary Data 2 | De-identified clinical presentation and vascular imaging summary. This worksheet provides de-identified summaries of clinical presentation and vascular imaging findings in patients with PAD. Direct identifiers, name-derived abbreviations, specific treatment locations and overly detailed narrative medical histories have been removed. Only research-relevant information related to PAD phenotype, vascular lesion extent, stenosis or occlusion, stents, bypass grafts and distal outflow tract status is retained. These data support interpretation of ABI-insensitive cases, perioperative case analyses and Supplementary Tables 2 and 6.

Supplementary Data 3 | Limb-level leave-one-subject-out cross-validation results. This worksheet provides limb-level subject-level LOOCV results, including sample ID, subject ID, side, true group, predicted group, classification correctness, PACT-DMS decision score and F1–F7 feature values. The F1–F7 values in Supplementary Data 3 represent model-input feature values used for PACT-DMS. For direction-oriented features, the sign reflects the predefined feature orientation and should not be interpreted as a raw physical count or area. These data provide source data for Fig. 3 and Fig. 4 of the main manuscript, Supplementary Fig. 12 and Supplementary Tables 4 and 6.

Supplementary Data 4 | Subject-level bilateral fusion results. This worksheet provides subject-level bilateral fusion results, including left-foot score, right-foot score, bilateral fusion score, true group, predicted group and classification correctness. These data support the classification performance of the bilateral fusion model, BiScore-related analyses, covariate sensitivity analysis and Supplementary Fig. 13.

Supplementary Data 5 | Vessel pulsation scoring. This worksheet provides vessel pulsation scoring records, including side, subject ID, observer 1 score and binary assessment, observer 2 score and binary assessment, reviewer score and binary assessment, and the final pulsation assessment. These data support the definition of Vessel Pulsation Strength (F3) in the main manuscript, the interobserver agreement analysis in Supplementary Fig. 10 and the traceability of F3 as a candidate dynamic vascular-behaviour feature.

**S**upplementary Data 6 | STARD 2015 reporting checklist. This worksheet provides the completed STARD 2015 reporting checklist for the diagnostic-accuracy component of the study, including item-level information on the study design, participant eligibility, index test, reference-standard assignment, blinding or independent review, diagnostic-performance measures, uncertainty estimation and participant flow. These data support transparent reporting of PACT-DMS as an investigational photoacoustic readout for PAD-related diagnostic discrimination and clarify where the corresponding information is reported in the main manuscript, Methods, Supplementary Information and Supplementary Data. This checklist is not intended to introduce additional experiments or diagnostic claims, but to document the reporting completeness of the current study.

Supplementary Data 7 | TRIPOD+AI reporting checklist. This worksheet provides the completed TRIPOD+AI reporting checklist for the model-development component of the study, including item-level information on the prediction objective, input features, classifier implementation, feature standardization, subject-level cross-validation, thresholding, data-leakage control, performance evaluation, robustness analyses, limitations, and data and code availability. These data support transparent reporting of the PACT-DMS diagnostic model and complement Supplementary Table 6, Supplementary Fig. 12 and Supplementary Fig. 13 by documenting how the model construction, validation and robustness analyses were reported. This checklist is not intended to establish an externally validated prediction model, but to provide reporting transparency for interpreting the current single-centre model-development results.

[1].Netter FH. Muscles, Arteries, and Nerves of Front of Ankle and Dorsum of Foot: Deeper Dissection; Dorsum of Foot: Deep Dissection. In: Atlas of Human Anatomy. 7th ed. Philadelphia, PA: Elsevier; 2019:522.

[2].Netter FH. Venous Drainage of Leg. In: Atlas of Human Anatomy. 7th ed. Philadelphia, PA: Elsevier; 2019:513.

[3]. Hamada N, Ikuta Y, Ikeda A. Arteries to the great and second toes based on three-dimensional analysis of 100 cadaveric feet. Surg Radiol Anat. 1993;15:187–192.

[4]. Sobotta J. Superficial nerves and veins of the dorsum of the foot. In: McMurrich JP, ed. Atlas and Text-book of Human Anatomy. Vol. 3, Vascular System, Lymphatic System, Nervous System and Sense Organs. Philadelphia, PA: W.B. Saunders; 1909. 721.
